# A human pan-genomic analysis reconfigures the genetic and epigenetic make up of facioscapulohumeral muscular dystrophy

**DOI:** 10.1101/2023.06.13.23291337

**Authors:** Valentina Salsi, Matteo Chiara, Sara Pini, Paweł Kuś, Lucia Ruggiero, Silvia Bonanno, Carmelo Rodolico, Stefano C. Previtali, Maria Grazia D’Angelo, Lorenzo Maggi, Diego Lopergolo, Marek Kimmel, Filippo M. Santorelli, Graziano Pesole, Rossella G. Tupler

## Abstract

Facioscapulohumeral muscular dystrophy (FSHD) is the only human disease associated with epigenetic changes at a macrosatellite array. Almost 95% of FSHD cases carry a reduced number (≤10) of tandem 3.3 kilobase repeats, termed D4Z4, on chromosome 4q35; remaining cases bear variants in chromatin remodeling factors, such as SMCHD1, DNMT3B, LRIF1. Reduced CpG methylation is used for the molecular diagnosis of FSHD, but D4Z4-like sequences dispersed in the genome can generate ambiguous results. By analyzing complete haplotype level assemblies from the T2T-CHM13 human genome and 86 haploid genomes from the human pangenome project, we uncovered the extensive number of D4Z4-like elements and their widespread inter- and intra-individual variability. An original analytical approach was developed to elucidate this previously unaccounted wealth of D4Z4-like elements and to analyze CpG methylation at D4Z4 in bisulfite-treated DNA from 29 FSHD index cases and 15 relatives. Integrated analysis of clinical phenotype, D4Z4 CpG methylation level and gene variants showed that low D4Z4 methylation was associated with variants in the *SMCHD1* gene, but not with the patients’ clinical phenotypes. This is the first study showing the relevance of the pangenome and T2T-CHM13 assemblies for investigating the genotype-phenotype correlation in genetic diseases. The extension and the variability of D4Z4-like elements scattered throughout the human genome and the inconsistent association of phenotypes with methylation profiles advocate for a critical revision of FSHD diagnostic tests based on D4Z4 CpG methylation assays and indicate that molecular investigations must be complemented by family studies for the proper interpretation of results.

## Introduction

Facioscapulohumeral muscular dystrophy (FSHD) is the third most common form of hereditary myopathy with an estimated prevalence from 1/8,000 to 1/20,000^1,2^. FSHD is the only hereditary human disease associated with reductions in copy number of a 3.3 kb macrosatellite^3^, termed D4Z4. In the general population, the size of the D4Z4 repeat varies between 11 and 150 units, whereas FSHD patients carry fewer than 11 repeats^4^. Each copy of the 3.3 kb D4Z4 repeat contains an open reading frame (ORF) with two homeobox domains, named *DUX4-*Like (*DUXL*)^5^, and two different classes of GC-rich repetitive DNA^6,7^: hhspm3, a member of a low copy human repeat family^8^, and LSau, a middle repetitive DNA family^8,9^. A tandemly arrayed D4Z4 repeat with 98% identity to the 4q35 array is located at 10q26^10^, but only alleles with reduced D4Z4 copy number on chromosome 4q35 have been associated with FSHD (Figure 1).

**Figure 1.**
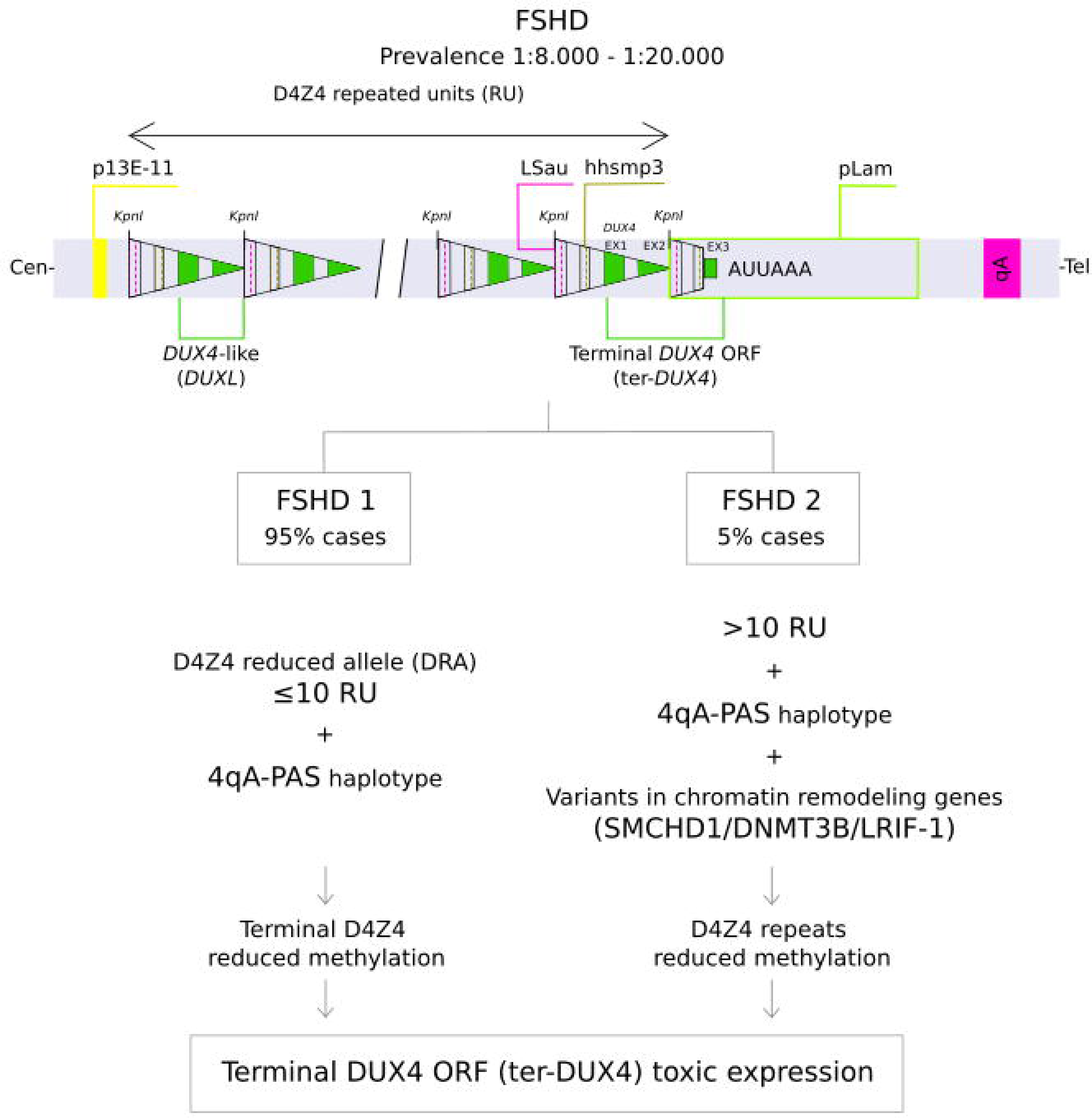
Scheme of the proposed models for FSHD1 and FSHD2 pathogenesis. Tandemly arrayed D4Z4 repeats (triangles) and the 4q35 haplotype elements are represented along with the molecular signatures proposed for FSHD1 and FSHD2 pathogenesis.

Two genetically distinct FSHD subtypes, FSHD1 and FSHD2, are currently described. In FSHD1 [OMIM #158900], the reduction of 4q35-D4Z4 repeats below a critical threshold (10 or fewer repeat units, RU)^11^ is thought to determine epigenetic alterations and the inappropriate expression of nearby genes leading to disease ^12–14^. In FSHD2 [OMIM #158901] affected individuals carry two D4Z4 arrays in the healthy range (>10 RU), but bear damaging heterozygous variants in chromatin remodeling factor genes, such as *SMCHD1* (for structural maintenance of chromosomes flexible hinge domain containing 1)^15^, *DNMT3B* (for methyltransferase 3B)^16^ and *LRIF1* (for ligand-dependent nuclear receptor interacting factor 1)^17^. It has thus been proposed that similar to the reduction of D4Z4 RU, the inactivation of these genes alters the epigenetic configuration of the D4Z4 array at 4q35 (Figure 1). Recent studies linked the molecular etiology of FSHD with the inappropriate expression of the DUX4 protein from the terminal D4Z4 repeat. Only a specific haplotype labelled 4qA-PAS is considered to be permissive for DUX4 protein expression, due to the presence of a 260-bp sequence, termed pLAM, which provides a 3’-terminal exon (Exon 3) and a polyadenylation signal (PAS) (AUUAAA) in juxtaposition to the terminal copy of *DUX4* ORF (hereafter ter-*DUX4*) on 4q^18,19^ (Figure S1). Although a similar arrangement is observed also on chr 10q26, at this locus the PAS signal is disrupted by a single nucleotide variant (AUCAAA), preventing the polyadenylation of the ter-*DUX4* mRNA. By analogy, 4qB, an alternative (to 4qA-PAS) haplotype at 4q (Figure S1), is considered not permissive for the expression of the DUX4 protein due to the lack of pLam. Based on these observations (D4Z4 reduced copy number or heterozygous variants in chromatin remodelers, associated with 4qA-PAS haplotype) a reduction of CpG methylation of D4Z4 has been proposed as a faithful indicator of anomalous *DUX4* gene expression and it is used as a proxy of disease status in the clinical setting^20^.

This model is in contrast with clinical and epidemiological data showing reduced penetrance and clinical variability in FSHD families^21–27^, as well as with observations obtained by adopting an articulate neurological assessment for the study of genotype-phenotype correlation in FSHD. This latter approach includes the phenotypic classification of probands and relatives by applying the Comprehensive Clinical Evaluation Form (CCEF)^28^, which beside quantifying the degree of motor disability^29^, classifies individuals on the basis of detailed phenotypic features which include parameters such as age at onset or site of disease onset beside the detailed description of muscle impairment. Four main descriptive categories have been created. They identify: (1) subjects presenting facial and scapular girdle muscle weakness typical of FSHD (category A, subcategories A1-A3), (2) subjects presenting an incomplete phenotype with muscle weakness limited to scapular girdle or facial muscles (category B, subcategories B1 and B2), (3) asymptomatic or healthy subjects (category C, subcategories C1 and C2), and (4) subjects with myopathic phenotypes presenting clinical features not consistent with FSHD canonical phenotype (category D, subcategories D1 and D2)^28^. The application of this methodology for the stratification of clinical phenotypes has permitted the quantification of the large phenotypic variability observed in individuals carrying a D4Z4 Reduced Allele (DRA) which is in contrast to the indication that a positive molecular test is the only determining aspect for FSHD diagnosis^30–37^. Studies also showed that the different categories are associated with different disease trajectories; in particular subjects assessed with a classical FSHD phenotype (Category A) display a steeper functional decline, than subjects with limited muscle impairment (Category B) ^31,35^. Large genotype-phenotype studies also highlighted differences between probands and relatives, including 30-50% of relatives remaining non-penetrant carriers. Consistent with these findings it has been assessed that DRA with 4-8 RU have the frequency of a common polymorphism (3% in the general population^25,38–40^), which is not compatible with the incidence of FSHD; moreover the DRA 4qA-PAS haplotype has a 2% frequency in the human population^39^. Finally, studies reported a wide variability in D4Z4 CpG methylation levels among FSHD index cases and FSHD families with no clear association between D4Z4 methylation status and disease manifestations or severity^32,41^.

Here, we report the results of comparative analyses of 86 haplotype-level assemblies from the Human Pangenome Project dataset (hereafter PGR)^42^ and the outcome of high-throughput CpG methylation assay of the D4Z4 repeat based on the T2T-CHM13 human genome assembly (hereafter T2T). Our study displays all the potential of complete, high-quality haplotype level genome assemblies for the analysis of D4Z4 arrays in human diseases, confirms the high frequency of the permissive 4qA-PAS haplotype, which is comparable to a common polymorphism and shows that across the 3.3 kb D4Z4 elements only two sequences are 4q/10q-specific and amenable for the study of methylation patterns in FSHD. This refined CpG methylation assay reveals that 4q/10q-specific reduced methylation correlates with the presence of damaging *SMCHD1* heterozygous variants, but not with the clinical status of FSHD2 cases and advocate revisions of previous findings based on the GRCh38 reference (hereafter hg38).

## Subjects and methods

### Participants

The study cohort included 26 subjects (8F, 18M) reported to the Miogen Laboratory of the University of Modena and Reggio Emilia for the diagnosis of FSHD^43^. These cases carried D4Z4 alleles with 10 or more repeats. In 8 cases the study was extended to one or both parents, for a total of 14 subjects (7F, 7M). Overall, 5 trios and 3 parent-child couples were analyzed. Four subjects carrying a 4U DRA were also included (2F, 2M). In three of these subjects (Family C individuals III-1 and III-3; Family 1252 subject 297/08^32^) D4Z4 methylation was previously assessed through the BSS assay using primer sets specific for the distal 4qA and 4qA-Long (4qA-L) D4Z4 repeat regions^32^.

The clinical phenotype was determined (Table S1) according to the Comprehensive Clinical Evaluation Form (CCEF), which evaluates the distribution and degree of motor impairment, age at onset of motor impairment and site of muscle weakness at onset, the presence of typical and atypical symptoms^24^. The CCEF has been developed by the Italian Clinical Network for FSHD to classify subjects on the basis of these clinical features. The CCEF classifies: 1) subjects presenting facial and scapular girdle muscle weakness typical of FSHD (category A, subcategories A1-A3), 2) subjects with muscle weakness limited to scapular girdle or facial muscles (category B subcategories B1, B2), 3) asymptomatic/healthy subjects (category C, subcategories C1, C2), 4) subjects with myopathic phenotype presenting clinical features not consistent with FSHD canonical phenotype (category D, subcategories D1, D2). Degree of motor impairment was also established by applying a standardized evaluation that generates the FSHD score ^29^.

Signed informed consent was obtained from all the subjects prior to the inclusion in the study.

### Bisulfite sequencing

Bisulfite sequence analysis was performed on high molecular weight genomic DNA obtained from peripheral frozen blood through phenol-chloroform extraction. To assess the methylation level at D4Z4 locus, specific primer sets were designed on hg38 genome assembly using MethPrimer tool^44^. No CpG sites were allowed in the primer sequence. The amplicon selection was based on following criteria: i) the sequences contain SNPs allowing to discriminate between the 4q and 10q D4Z4 repeats; and ii) include the maximum number of CpG sites possible. Amplicons were representative of the whole D4Z4 array and included functional domains such as the D4Z4 binding element (DBE)^12^. Table S2 and Table S3, Figure S2 summarize the characteristics of the four selected primer sets. Specific Illumina adapters were added to each set of primers.

Bisulfite conversion was performed on 1 μg of genomic DNA by using the EpiTec Bisulfite Kit (Cat N°59104 QIAGEN) following the manufacturer’s protocol.

PCR amplification of both converted and non-converted DNA was performed using the four selected primer pairs (Table S2). For every subject we generated two pools of amplicons: one from bisulfite converted (BSC), one from non-converted DNA. Illumina paired-end sequencing was then performed by Eurofins genomics on the amplicon pools for a total of 36 Mb (60K read pairs per amplicon with 2 x 300 bp read mode).

### Bioinformatics analyses

#### Identification and characterization of D4Z4-like sequences and D4Z4 associated elements in human genome assemblies

The complete sequence of a reference D4Z4 element was obtained from the hg38 assembly in UCSC genome browser (https://genome.ucsc.edu/). Sequence similarity searches based on the BLAST program^45^ were performed to annotate D4Z4-like elements and functional elements of the D4Z4 array (see below), in the hg38 reference assembly of the human genome [<u>GRCh38.p14 - hg38 - Genome - Assembly - NCBI</u>], the T2T assembly [<u>T2T-CHM13v1.1 - Genome - Assembly - NCBI</u>], and 86 distinct haplotype-level assemblies from the PGR, as available from https://github.com/human-pangenomics/HPP_Year1_Assemblies.

Results of BLAST sequences similarity searches were stored in simple tabular format and processed by custom Perl scripts. A similarity threshold of 85% and an alignment length threshold of 300 aligned or more aligned residues were used to define D4Z4-like elements. Matches were binned in 5 bins according to their size (<500bp; <1000bp; <1500bp; <2500bp; and <3300bp). Matches of size in-between 3200 and 3300 bp were considered to provide a complete representation of a D4Z4 macrosatellite (full match).

D4Z4 arrays were annotated based on the terminal repeat for the presence of qA or qB sequence variants; presence/absence of the pLam sequence and integrity of the AUUAAA polyadenylation signal (PAS). qA/B has been annotated based on the sequence of the respective probes used for the FSHD diagnostic protocol; pLam has been annotated based on the sequence reported in UCSC genome browser (https://genome.ucsc.edu/)

(Supplemental Material and Methods).

#### Similarity-based clustering of D4Z4 like elements and terminal DUX4 coding sequences

D4Z4-like sequences of 3200 or more bp in size (full matches) were extracted from their respective genome assemblies and aligned with Kalign^46^. Conserved alignment blocks were extracted with Gblocks^47^, using the following parameter: Minimum Length Of A Block: 3, Allowed Gap Positions: none. A sequence similarity matrix was computed by means of the EMBOSS distmat program^48^, using the Tajima-Nei correction for multiple substitutions^49^. Phenetic clustering was performed by using the NJ algorithm, as implemented by hclust() function from the cluster package in R^50^. Six distinct groups with high sequence identity were defined for the D4Z4 complete elements. D4Z4 from T2T were used to anchor/assign each group to a chromosome.

The same method was applied to cluster ter-*DUX4* coding sequences by sequence similarity. Terminal repeats were identified based on the presence of a qA or qB element *in cis* and within 15 Kb from a complete *DUX4* ORF and tentatively assigned to chr 4 or 10 based on sequence similarity.

#### In-silico identification of target regions of primer sets in hg38 and T2T genome assemblies

Potential target regions of primer sets in the hg38 and T2T genome assemblies were determined by a custom Perl script (Table S4). Sequence similarity searches of primer sequences on the hg38 and T2T reference assemblies were performed by blastn, with default parameters^45^. All matches of 17 bp or longer and with at most 1 mismatch and all perfect matches of 16 bp or longer were recorded. Genomic coordinates were cross-referenced, and potential target genomic regions were identified as those spanned by a 5’ and 3’ primer pairs, in the correct orientation, and within a distance >50 bp and <600 bp. The same procedure was applied for the identification of potential primer target regions in the BSC space, in this case both the genome and the primer sequences were converted *in-silico*.

### Statistical analyses

Distribution of values were compared by applying the non-parametric, two tailed Kolmogorov Smirnoff test, as implemented by the ks.test() function from the stats package of the R programming language.

### Analysis of bisulfite converted reads

#### Quality control and determination of target regions

Reads’ quality was assessed by Fastqc^51,52^. Individual reports were merged by MultiQC^53^ and quality metrics were visually inspected.

Non-converted amplicons’ reads were aligned to T2T and hg38 human genome assemblies using Bowtie2^54^. Sample-level coverage profiles were computed by bedtools genomecov^55^. Genomic regions covered by 100 or more reads in at least 50% of samples were considered for subsequent analyses. By this approach a total of 172 and 57 candidate target regions were identified in the T2T and hg38 assemblies, respectively. 50/57 and 169/172 had at least a high similarity match (17 bp with at most 1 mismatch) to one or more primer sequences.

Highly similar candidate regions were collapsed to provide a non-redundant representation of repetitive sequences and allow non-ambiguous mapping of BSC reads. Different sequence identity thresholds were tested through a titration analysis to identify the ideal sequence similarity threshold. The total number of non-ambiguously mapped BSC reads and the total number of CpGs covered by at least 10 BSC reads in at least 50% of samples (testable CpGs) were recorded (Table 1). Considerations based on the number and cumulative size of genomic regions that were merged, and on the number of testable CpGs, prompted us to select 95% identity as the most appropriate threshold. In conclusion, by comparing hg38 and T2T-based analyses of non-converted reads, we defined a total of 60 and 9 distinct consensus groups (groups of sequences GS) respectively (Table S5 and Table S6). For every target region the consensus sequence was determined by majority rule consensus, by applying the cons EMBOSS program^48^. These consensus sequences were used for the further analysis of methyl-seq data.

**Table 1.** Titration analysis for the identification of the optimal sequence similarity threshold.

| Identity-level (%) | # regions | size (Kb) | uniquely mapped<br>BSC reads (%) | # testable<br>CpGs |
| --- | --- | --- | --- | --- |
| 99 | 145 | 67,41 | 5,43% | 237 |
| 98 | 103 | 59,13 | 12,13% | 411 |
| 97 | 87 | 54,01 | 26,74% | 873 |
| 96 | 73 | 46,14 | 32,47% | 997 |
| *95* | *60* | *40,03* | *49,17%* | *1176* |
| 94 | 56 | 38,71 | 51,01% | 1083 |
| 93 | 53 | 37,55 | 53,43% | 891 |
| 92 | 46 | 36,49 | 58,47% | 766 |
| 91 | 45 | 33,02 | 72,71% | 637 |
| 90 | 42 | 32,75 | 75,41% | 451 |

Identity-level (%): sequence identity threshold. #regions: number of distinct regions defined by the threshold. size (Kb): total size of the regions in Kb. uniquely mapped BSC reads (%): percentage of uniquely mapping BSC reads. # testable CpGs: total number of distinct CpGs for which more than 10 reads were available for more than 25 subjects. Italic characters indicate the selected sequence similarity threshold.

#### Assessment of methylation levels

BSC reads were aligned to consensus target region sequences and methylation levels were determined by Bismark^56^ +Bowtie2. The bismark_methylation_extractor tool was applied to compute CpG methylation levels. Only CpGs covered by more than 10 reads in at least 25 patients were considered for the delineation of methylation profiles.

#### *SMCHD1* variants analysis

FSHD2 probands are routinely tested for the presence of *SMCHD1*, *DNMT3B*, *LRIF1* variants. We collected the identified *SMCHD1* variants in the cohort and performed a reannotation on GRCh37 genome assembly using wAnnovar^57^ (https://wannovar.wglab.org/; the analysis was performed on the 27.9.22) and filtered. We excluded intron variants and considered only exonic (missense, stop or frameshift variants) and splicing variants with a GnomAD exome all MAF < 10^-4^ (GnomAD frequency for PM2 pathogenic moderate rule in Varsome)^58^. Among the filtered variants we then excluded the ones which were benign and likely benign according to Varsome ACMG (American College of Medical Genetics and Genomics) prediction^59,60^ (https://varsome.com/; the analysis was performed on 27.9.22). As *SMCHD1* gene function has not been thoroughly investigated yet, we decided not to filter out the variants predicted as VUS (Variants of Uncertain Significance). Finally, variant validation through Sanger sequencing was performed on probands and on relatives (Table S7).

## Results

### T2T and PGR reveal the extended variability of D4Z4 repeats throughout the human genome

The extent of D4Z4-like repeat elements in the human genome was investigated by analyzing T2T and 86 haplotype-level genome assemblies from the PGR^42^.

Our analyses uncovered hundreds of “complete” (>3200 bp) as well as partial (300 – 3200 bp) D4Z4-like sequences (hereafter D4Z4-l), irrespective of the reported geographic origin of the subjects (Table S8). The cumulative size of D4Z4-l spanned in-between 700 Kb to 1.5 Mb of sequence in the haplotype-level assemblies of the 43 subjects from the PGR and accounted for approximately 1.2 Mb of sequence in T2T. D4Z4-l were scattered through several chromosomes in T2T and accounted for hundreds of kilobases of sequences on chr 1, chr 4, chr 10, chr 15, chr 21 and chr 22 (Figure 2A). Notably, in hg38 only 86.5 Kb of D4Z4-l were observed, representing a more than 10-fold reduction compared with T2T and PGR (Figure 2B). In hg38, D4Z4-l were prevalently associated with the long arms of chr 4 and chr 10 (Figure 2A) where they are arranged as two large arrays of 8 and 10 complete D4Z4 repeats, respectively. PGR and T2T instead harbored a large number (minimum 285, maximum 626, Table S8) of “incomplete” D4Z4-l repeats of < 2.5 Kb in size, which are lacking in hg38. Conversely (Figure 2B), the number of complete D4Z4 elements included in hg38 was within the range of variability observed in T2T and PGR (26 to 121). Interestingly, subjects from AFR (African) ancestry were associated with a higher variability in D4Z4-l copy numbers (Kolmogorov Smirnoff test p-value p-value = 0.0006403), while a narrower range was observed in individuals of EAS (Eastern Asia) and AMR (South America) ancestry (Figure S3).

**Figure 2.**
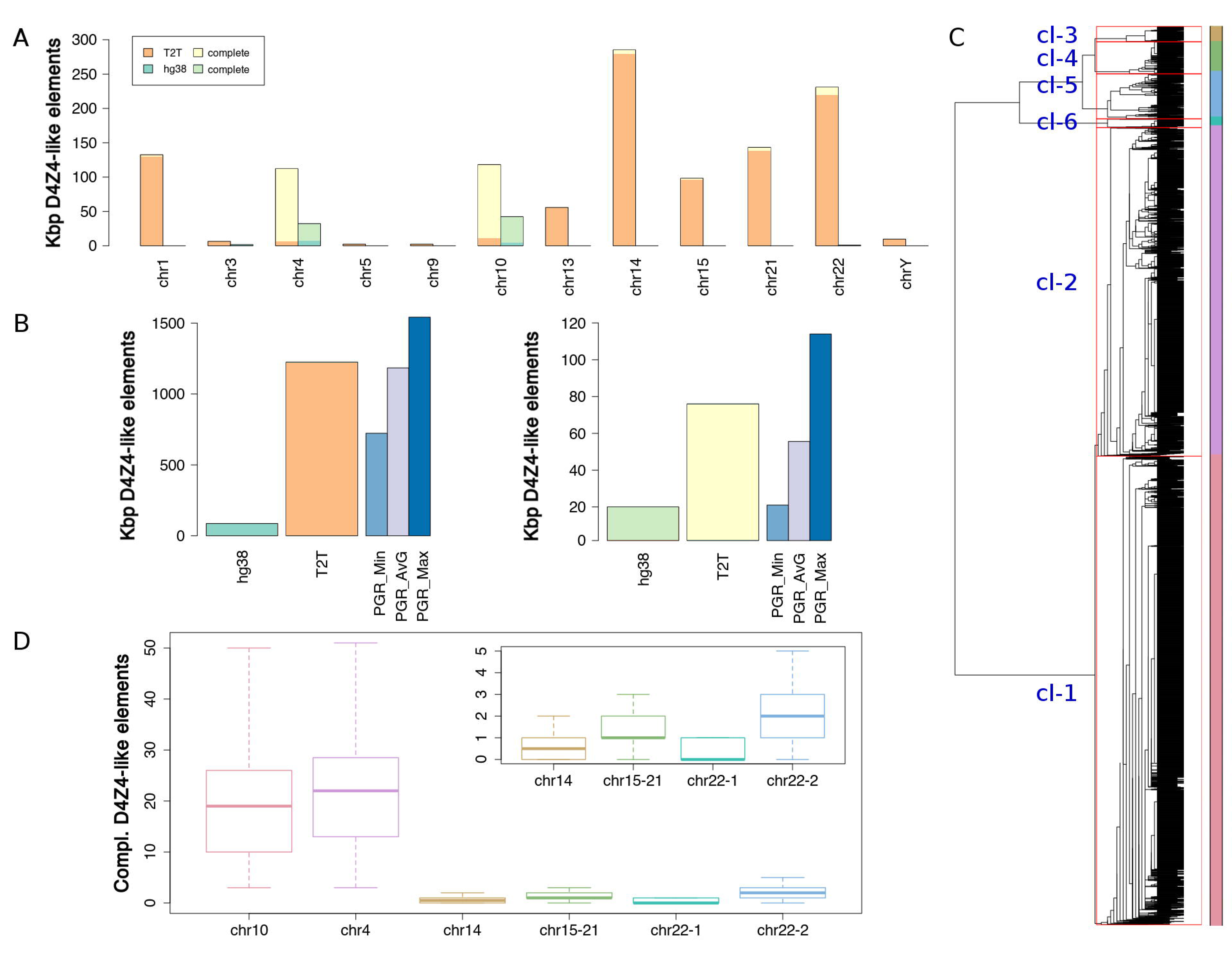
Stratification and variability of D4Z4-l elements. (A) D4Z4-l elements in hg38 and T2T. Cumulative size per chromosome of D4Z4-l elements. Only chromosomes for which at least 1 Kb of D4Z4-l sequence was identified, in any of the 2 assemblies, are reported. Size (in Kb) is shown on the Y axis. Different colors are used for T2T and hg38 and to mark complete D4Z4-l (see legend). (B) Cumulative size (in Kb) of D4Z4-l elements in the hg38, T2T and PGR haplotype level assemblies. Left: cumulative size of D4Z4-l elements. Right: cumulative size of full D4Z4-l elements. Data is displayed in the form of a barplot. Values are on the Y axis, labels on the X axis. For the human pangenome haplotype level assemblies, 3 distinct bars are used to indicate the minimum, maximum and average values. Color codes according to the legend. (C) Dendrogram of complete D4Z4-l elements. Red rectangles and color codes are used to delineate the 5 main groups of D4Z4-l elements, as identified by sequence similarity-based clustering. (D) Total estimated number of complete D4Z4 elements per sequence similarity group. Distributions of values are displayed in the form of a boxplot, with groups indicated on the X axis and copy numbers on the Y axis. The top-right panel provides zoom on groups chr 14, chr 15-21, chr 22-1 and chr 22-2.

Figure 2C shows the variability and the distribution of complete D4Z4-l elements. Six different groups were defined on the basis of sequence identity levels. D4Z4 from T2T were used to anchor/assign each group to a chromosome. The largest clusters (Table 2) and the majority of the sequences (87%) were assigned to either chr 4 (cluster-2) or chr 10 (cluster-1); this notwithstanding, complete D4Z4 elements were also observed on chr 14 (cluster-3), chr 22 (cluster-5 and cluster-6), chr 15 and chr 21 (cluster-4). The number of complete D4Z4 elements assigned to distinct clusters in the 86 PGR haplotypes is summarized in Figure 2D and Table 2. Chr 4 and chr 10 were associated with a median number of 22 and 19 repeats, respectively. The difference was statistically significant according to a Kolmogorov Smirnov test (p-value: 0.02518). Complete D4Z4 repeats on chr 10 displayed lower variability compared with those on chr 4. Substitution rates estimates at haplotype levels suggested a high inter-individual variability, with rates ranging from 0.164 to 0.4312 (average of 0.245) substitutions for 100 bp for chr 4 and from 0.015 to 0.21 (average of 0.13) for chr 10 (Figure S4 and Figure S5). Interestingly, as shown in Table 2, more than 75% of the assemblies carried D4Z4-l assigned to cluster-6 (chr 22); whereas less than 50% of the haplotype assemblies in the PGR had one sequence assigned to cluster-3 (chr 14), cluster-4 (chr 15-21) and cluster-5 (chr 22).

**Table 2.** Characterization of complete D4Z4 elements clusters.

| Cluster | Chromosome | % sequences | Median<br>number of<br>complete<br>repeats (per<br>haplotype) | Average<br>substitution<br>rate per 100<br>bp | Substitution<br>rates estimates<br>at haplotype<br>level |
| --- | --- | --- | --- | --- | --- |
| 1 | Chr 10 | 87% | 19 | 0.13 | 0.015-0.21 |
| 2 | Chr 4 |  | 22 | 0.245 | 0.164-0.4312 |
| 3 | Chr 14 | 2.3% | 0.5 | 1.73 | NA |
| 4 | Chr 15, Chr 21 | 3.1% | 1 | 1.81 | NA |
| 5 | Chr 22-1 | 1.5%+6.1% | 0 | 0.798 | NA |
| 6 | Chr 22-2 |  | 2 | 1.24 | NA |

### The pangenome assemblies show the high frequency of the 4qA-PAS haplotype associated with FSHD in the world population

The permissive DRA 4qA-PAS haplotype has been proposed to cause FSHD. However, we previously reported a frequency of 2% for this haplotype^38^, which is not compatible with the prevalence of the disease^61^. We thus analyzed the T2T and PGR to investigate the prevalence of this molecular signature in other populations and to study the conservation and the integrity of ter-*DUX4*.

The pLam and telomeric qA and qB sequences (Supplemental Material and Methods) were used to identify and classify terminal D4Z4 repeat in PGR, T2T and hg38. A total of 172 D4Z4 terminal repeats were identified (Table S9) and tentatively assigned to chr 4 or chr 10 based on sequence similarity profiles (as described by the clustering of the D4Z4-l sequences in Figure 2C). Haplotypes with inconsistent annotations (2 or more qA/qB probes assigned to the same chromosome and none to the other chromosome) were not included in subsequent analysis^62,63^. By this approach the complete set of alleles at D4Z4 repeat loci at chr 4 and chr 10 was determined for 72 haplotypes from PGR (Table S9). Patterns of conservation of ter-*DUX4* sequences were investigated. Figure 3A shows that ter-*DUX4* assigned to the 10q D4Z4 terminal repeat are found to be highly similar and formed a single cluster; while 4q ter-*DUX4* are more heterogeneous and were partitioned in 3 groups (Figure 3A-B). Interestingly, as shown in Figure 3C, in chr 10 and chr4_group 2 ter-*DUX4* displayed lower levels of variability (average 0.061 subs per 100 bp) compared to both chr4_group 3 (average 0.12 subs per 100 bp) and chr4_group 1 (average 0.142 subs per 100 bp); Figure 3A also shows that the most variable sequences (chr4_group 1 and chr4_group 3) were preferentially associated with qB alleles. All ter-*DUX4* sequences were found to be intact, and none was associated with frameshifts or premature stop codons. Consistent with previous observations^18,19^, all the alleles classified as qB did not carry a pLam and were more proximal to the ter-*DUX4* compared to qA (Figure 3D-G). In the same way, (Figure 3E-F) the vast majority of alleles classified as qA (112/119) were associated with a pLam both on chr 4 (41/47) and chr 10 (71/72). Only terminal 4q pLam had valid PAS^64^; while in pLam assigned to 10q the PAS sequences were disrupted by a single nucleotide substitution (AUUAAA→AUCAAA). Perfect conservation of PAS/PAS-like elements was observed both on 4q and 10q terminal D4Z4 repeats. One qB haplotype was assigned to 10q (Table 3).

**Figure 3.**
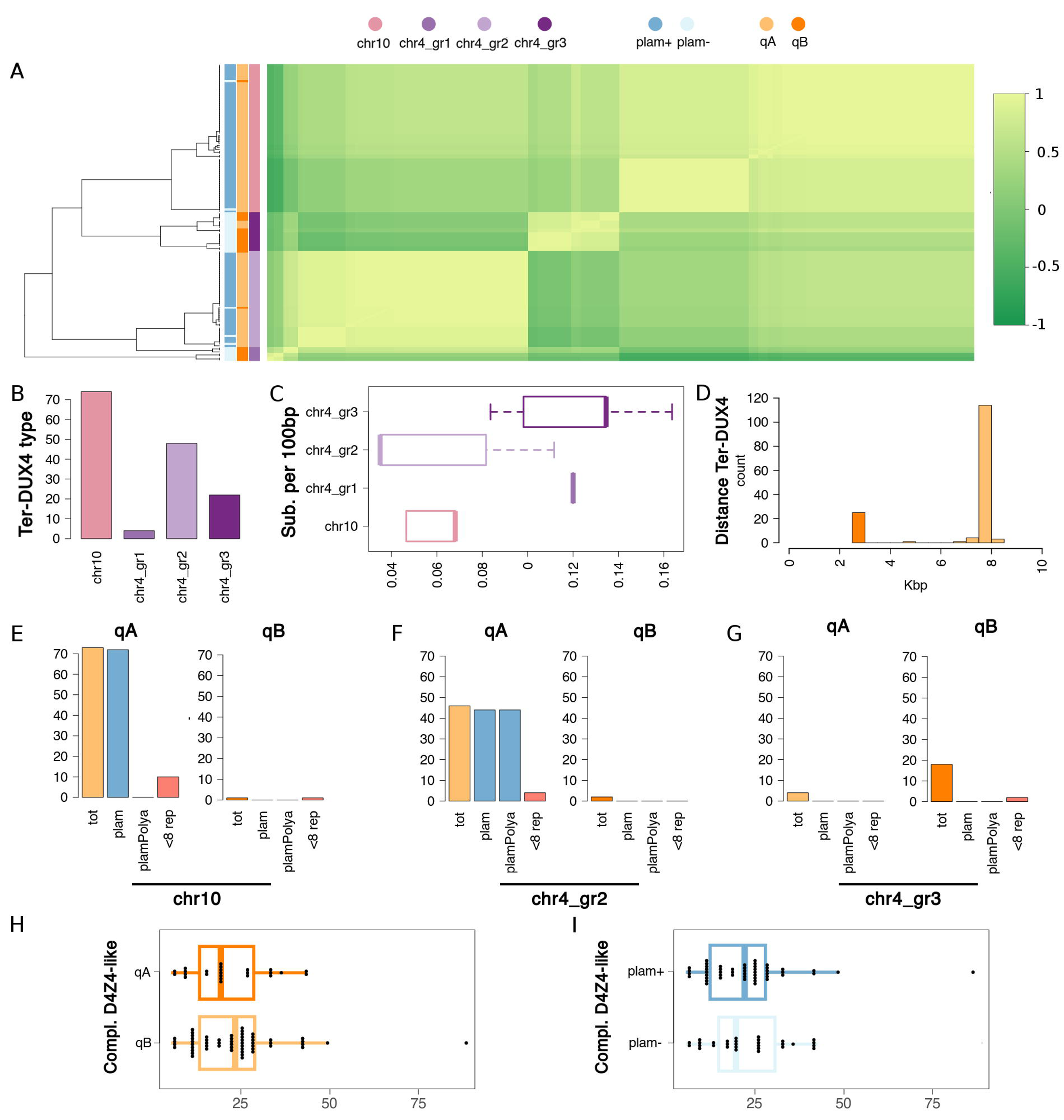
Characterization of D4Z4 alleles (A) Heatmap of ter-*DUX4*. Scaled identity levels are displayed by the green color gradient on the right. The dendrogram on the left shows clustering of ter-*DUX4* based on sequence identity. Colored vertical bars are used to indicate: presence/absence of pLam; classification as qA or qB; sequence similarity cluster (see top). Color codes are explained directly under each bar. (B) Numerosity of sequence similarity clusters. The barplot shows (y-axis) the total number of ter-*DUX4* in each cluster (x-axis). (C) Boxplot of substitution rates. Substitution rates (substitutions by 100 bp) by cluster. Distribution of values are shown as a boxplot. Clusters are on the y-axis, values on the x-axis. (D) Distance of qA and qB elements from ter-*DUX4*. Data are represented in the form of a histogram. Distances (in Kb) are indicated on the x-axis. qA and qB elements are marked with different colors (see legend). (E-G) Annotation of terminal D4Z4 repeats. Two barplots are displayed for the 3 (out of 4) clusters to which 5 or more sequences have been assigned. Each barplot reports, for qA and qB alleles respectively: the total count (tot); the number pLam-like elements (pLam); the number pLam-like elements with a valid PAS (plamPolyA); the total number of haplotypes with 8 or less D4Z4 repeats. Values are indicated on the y-axis, labels on the x-axis. Distinct sequence similarity clusters are indicated by a corresponding label directly under each plot. (G-H) Total number of complete D4Z4-l elements in H) qA and qB haplotypes; I) haplotypes with (pLam+) and without (pLam-). Distribution of values are displayed in the form of a boxplot, with values on the X-axis and groups on the Y axis. Color codes are consistent throughout Figure 2.

**Table 3.**
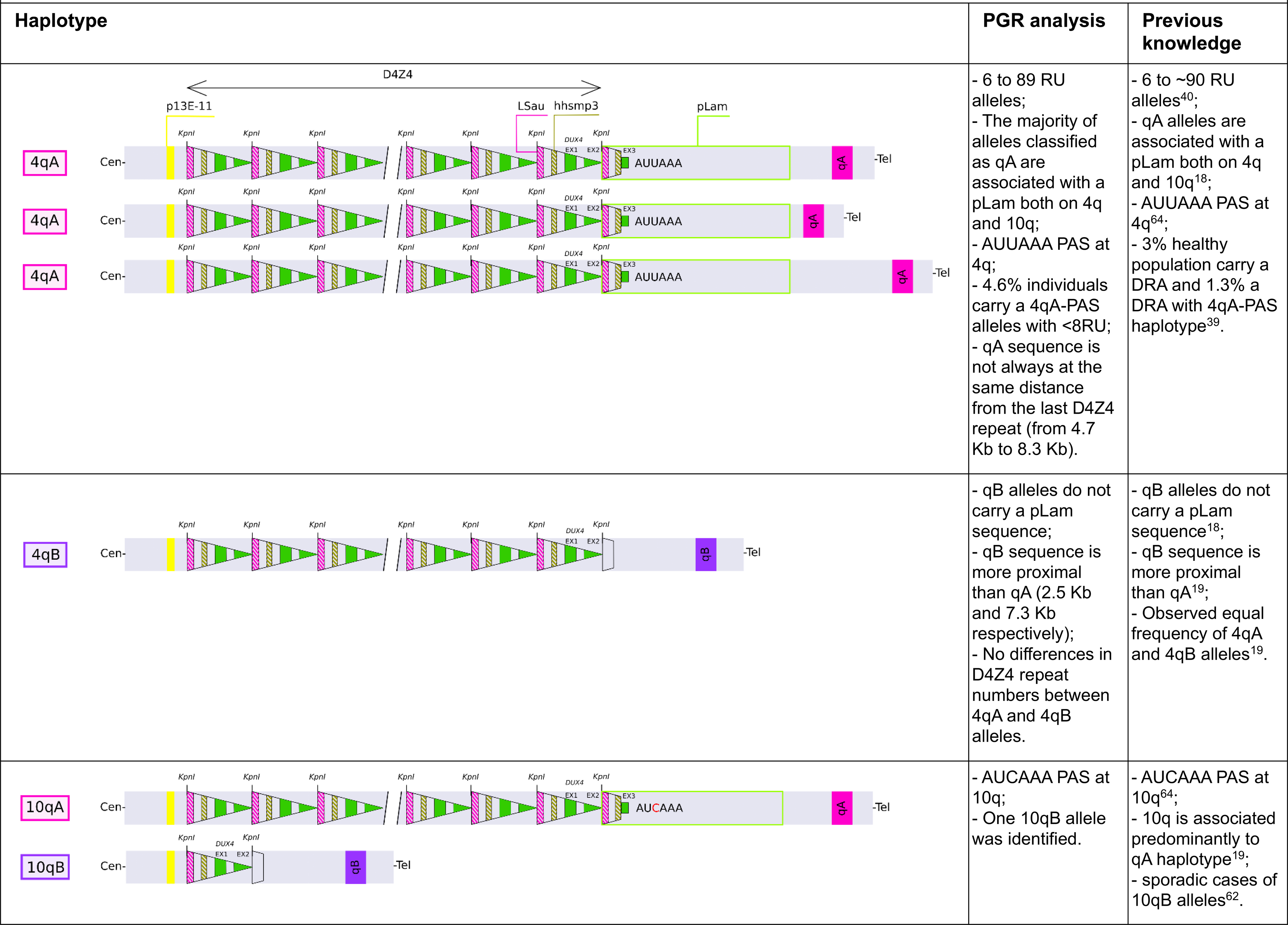
-Comparison between PGR analysis results and previous knowledge.

Remarkably, in line with our previous observations^38–40^ (Table 3), 4/86 haplotypes (4.6%) carried D4Z4 alleles with 6-8 RU and 4qA-PAS (Figure 3F). None of these haplotypes carried disruptive variants in the ter-*DUX4* or in the pLam sequence element.

No statistically significant difference in the total number of chr 4 D4Z4 repeats was observed when qA/qB alleles pLam+/pLam-alleles were compared (Figure 3H-I), suggesting that alleles that are not permissive for the expression of the ter-*DUX4* are not associated with significant differences in D4Z4 copy numbers compared with the permissive qA-PAS haplotype.

### D4Z4 hypomethylation correlates with *SMCHD1* damaging variants but not with FSHD clinical phenotype

Results shown above uncover all the heterogeneity of D4Z4 repeats in the human genome, including the presence of qA D4Z4 alleles with 8 repeats or fewer, and raise potential concerns about the reliability of methylation/diagnostic tests based on the hg38. As bisulfite treated DNA sequencing at targeted regions is commonly used to diagnose FSHD^65^, the uncovered variability of D4Z4-l sequences prompted us to investigate D4Z4 CpG methylation comparing the hg38 and the T2T.

A set of primers spanning the entire length of the D4Z4 repeat and a total of 126 CpGs (Figure S2 and Table S3) were designed based on hg38 assembly to recapitulate previous findings in the field^41^. This experimental design was used to assess a carefully selected cohort of 26 index cases referred for FSHD2 diagnosis, and 14 relatives all carrying 10 or more RU. All the participants assayed were searched for damaging variants in the chromatin remodelers *SMCHD1, DNMT3B* and *LRIF1*. In addition, 3 FSHD1 index cases and 1 relative carrying 4 RU were studied.

Amplicon targeted regions were sequenced both in non-converted and in BSC DNA samples. This approach was undertaken: i. to test the 4q/10q-specificity of the primer sets commonly used and designed based on hg38 assembly^41^, ii. to facilitate the alignment of short BSC derived reads. Both non-converted and BSC NGS data were used for sequence alignment based on hg38 and T2T reference genomes (Table S4).

A total of 77% and 99.43% non-converted short reads mapped to hg38 and T2T respectively. Moreover, the majority of BSC reads (62% for T2T and 54% for hg38) did not align at 4q primer target regions, potentially suggesting “off target” sequences, and/or high levels of heterogeneity in methylation levels. To identify genomic regions targeted by our assay in an unbiased manner, genomic intervals covered by 100 or more non-converted reads in at least 50% of the samples were recorded. A total of 57 and 172 genomic regions in hg38 and T2T displayed these remarkable levels of coverage. Notably, 50/57 and 169/172 of the target regions delineated by coverage analyses had at least a high similarity match (17 bp with at most 1 mismatch) to one or more primer sequences. Based on these observations sequences above 95% sequence similarity were collapsed to prevent inconsistent mapping of BSC short reads across highly similar D4Z4-l elements. By this approach, we defined a total of 60 and 9 distinct consensus groups (groups of sequences, hereafter GS) in T2T and hg38 respectively (Table S5 and Table S6). Consensus sequences were computed for each GS and used for the further analysis of methyl-seq data.

This *ad-hoc* bioinformatics workflow revealed remarkable differences in the completeness, number, and breadth of testable CpGs when the GSs derived from hg38 and T2T were compared: hg38 125 CpG; T2T 938 CpG. Global methylation patterns were summarized in the form of a heatmap (Figure S6 and Figure S7). In the T2T-based analysis a total of 4 distinct GS including 81 distinct CpGs had no missing data and complete methylation profiles across all samples, as shown in Figure 4 and Table 4: GS1 (primer D6): 28 CpGs; GS2 (primer D1): 32 CpGs; GS5 (chr 13-chr 15): 16 CpGs and GS6 (chr 13-chr 15): 5 CpGs). Notably, only GS1 and GS2 included D4Z4 5’ 4q/10q-specific regions exclusively, for a total of 60 CpGs, while GS5 and GS6 mapped in D4Z4-l repeats on chr 13 and 15. The wide-range distributions of CpG methylation levels, as displayed in Figure S8, highlight a high variability at regions GS1 and GS2, while off-target regions GS5 and GS6 have high methylation levels, with a narrower distribution. Figure S9 shows a general overview of the CpG methylation patterns observed in all the 60 distinct GSs defined in our analysis. Taken together these findings indicate that a significant proportion of the BSC reads recovered by our targeted high throughput assay derive from off-target, hyper-methylated D4Z4-l regions. We also observed that when hg38 is considered as reference, the criteria we established for the T2T-based analysis (namely no missing data and complete methylation profiles across all samples) identified only a single group, GS3 (primer D1) including 26 CpGs (Figure 4, Figure 5 and Table 4). In the light of these observations, methylation profiles inferred from T2T were considered more informative and were selected for subsequent analyses. Only primer D1 had complete data both in T2T and hg38 (Table 4); however, CpG methylation levels estimated on hg38 were slightly more elevated compared to those observed on T2T at the equivalent target region (Figure S10, median values: 49.13 hg38, 43.75 T2T). An observation that might be consistent with discrepancies in the mapping of BSC reads on different genome assemblies.

**Figure 4.**
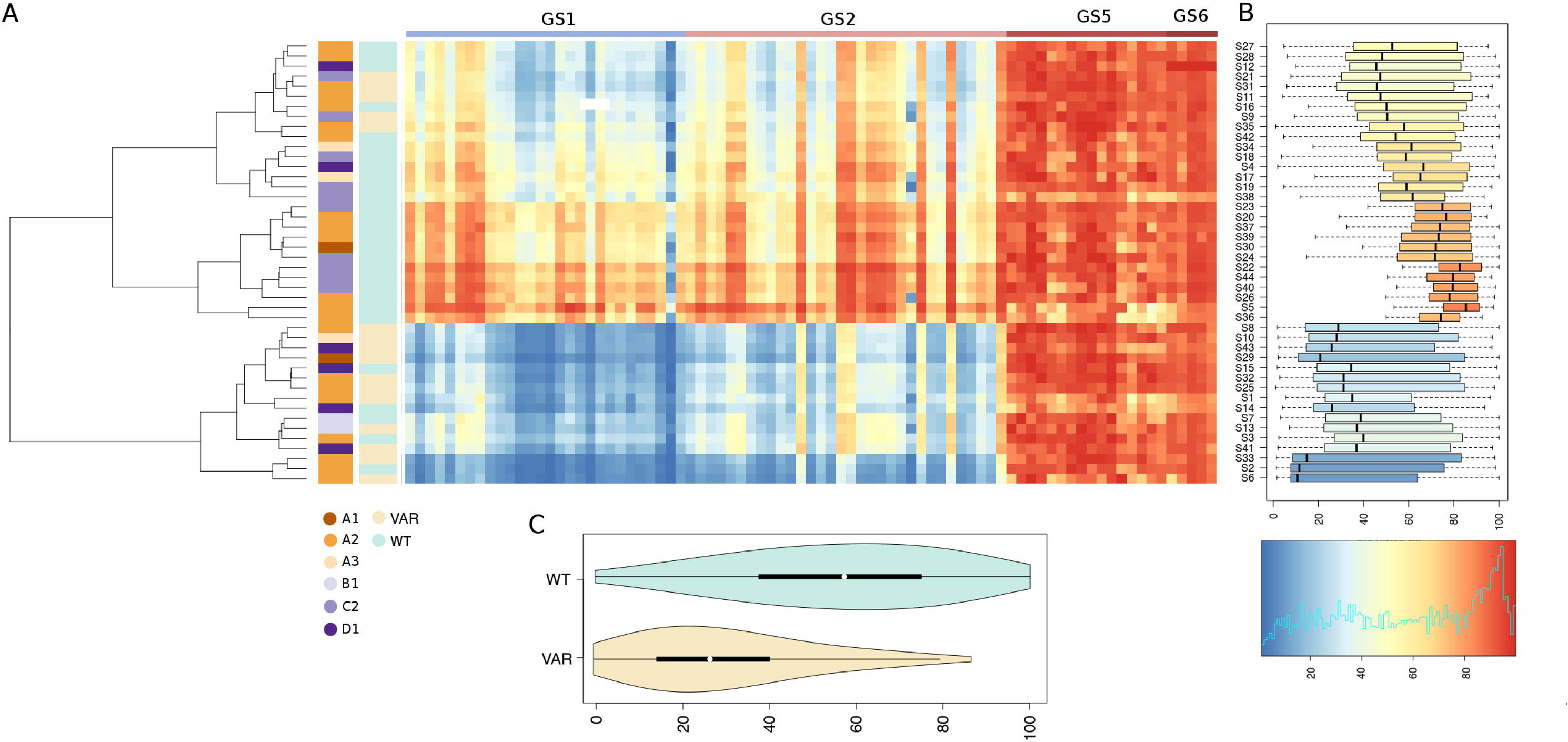
CpG Methylation profiles of cohort subjects inferred on T2T. A total of 82 CpGs for which complete data are available for all the study subjects are shown. (A) Heatmap of % CpG methylation: each row represents the CpG methylation pattern of a single individual. Columns represent the methylation pattern of each analyzed CpG. The dendrogram shows the clustering of the subjects based on the observed methylation profiles. Colored vertical bars are used to display CCEF clinical category status, and the presence/absence of deleterious genomic variants in *SMCHD1*. Color codes are illustrated directly under each bar. The colored bar at the top demarcates each of the 4 distinct 95% sequence identity genomic region clusters with complete data for all the subjects. (B) Boxplots of methylation levels distributions by subject. The color-scale used for the heatmap and the boxplot is shown at the bottom. (C) Violin-plot of average methylation levels, restricted to CpGs in regions GS1 and GS6, in subjects with/without deleterious genetic variants in *SMCHD1*.

**Figure 5.**
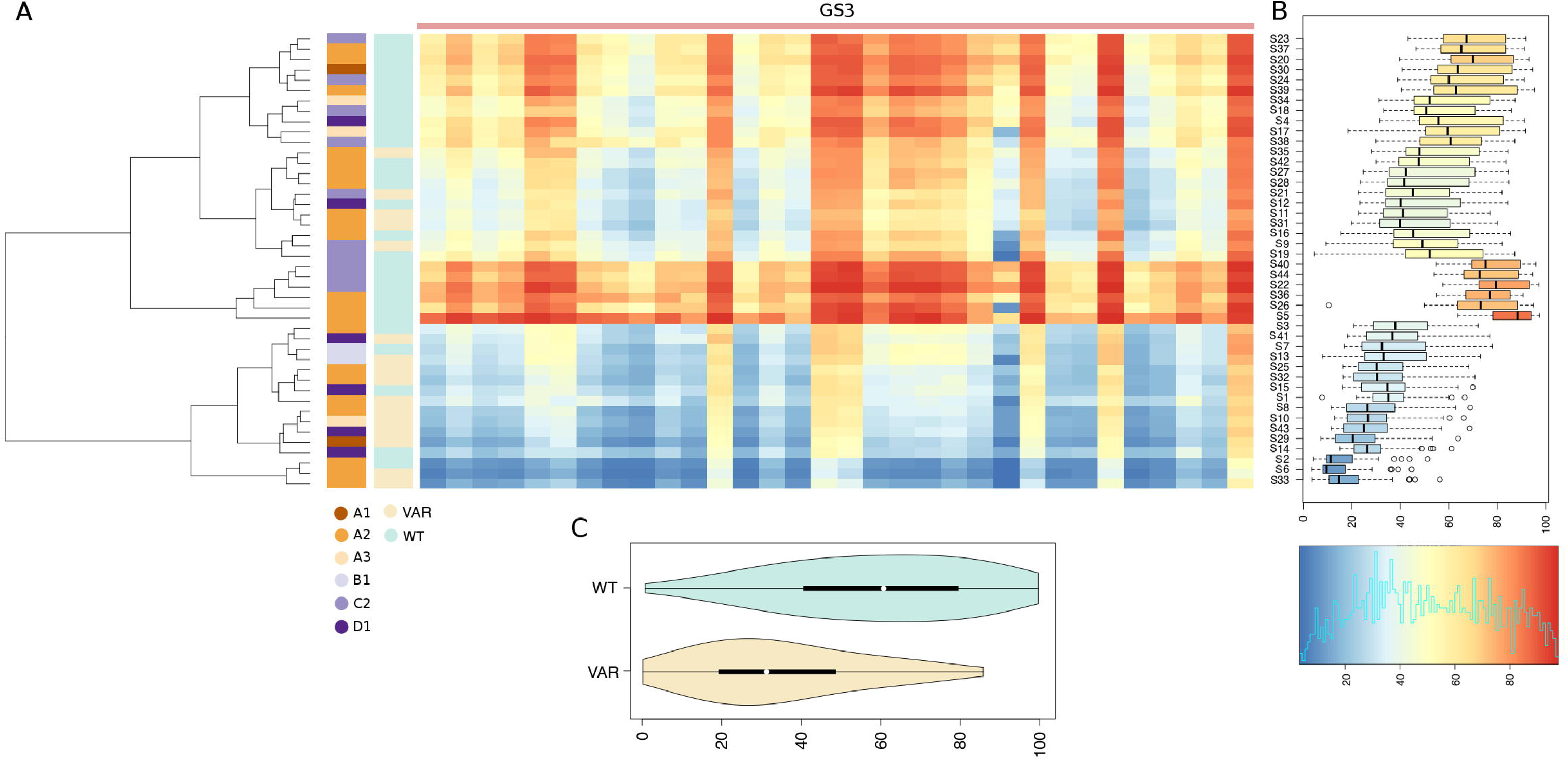
Methylation profiles of cohort subjects inferred on hg38. A total of 32 distinct CpGs, for which complete data are available for all the study subjects, are displayed. (A) Heatmap of % CpG methylation: individuals are reported on the rows and CpGs on the columns. The dendrogram shows a clustering of the subjects based on the observed methylation profiles. Colored vertical bars are used to display CCEF grades, and the presence/absence of deleterious genomic variants in *SMCHD1*. Color codes are illustrated directly under each bar. The colored bar at the top demarcates each of the 4 distinct 95% sequence identity genomic regions clusters with complete data for all the subjects. (B) Boxplots of methylation levels distributions by subject. The color-scale used for the heatmap and the boxplot is shown at the bottom. (C) Violin-plot of average methylation levels in subjects with/without deleterious genetic variants in *SMCHD1*. Only CpGs in primer-targeted regions are considered.

**Table 4:** GS containing CpGs covered by at least 10 BSC reads in at least 50% of samples and no missing data across all samples.

| Assembly | Group | Chromosome | N° CpGs |
| --- | --- | --- | --- |
| T2T | GS1 | Chr 4, Chr 10 | 28 |
|  | GS2 |  | 32 |
|  | GS5 | Chr 13, Chr 15 | 16 |
|  | GS6 |  | 5 |
| hg38 | GS3 | Chr 4, Chr 10 | 26 |

Further, our analysis revealed that 4q/10q-specific methylation patterns at GS1 and GS2 stratified samples into 3 groups: low methylation (average CpG methylation 24%), intermediate methylation (average CpG methylation 48%), and high methylation (average CpG methylation 71%). The methylation pattern of GS5 and GS6 was highly uniform across all samples, with an average of 89%.

Interestingly, the observed methylation profiles did not correlate with the disease status. As shown in Figure 6A, of 16 samples presenting low methylation 10 (62.5%) derived from cases presenting the classical FSHD features (CCEF category A), 6 (37,5%) derived from cases presenting incomplete (2, CCEF clinical Category B) or complex (4, CCEF clinical category D) phenotypes. Of 16 samples, presenting intermediate methylation 9 (56.3%) derived from cases assessed as clinical category A. Of 12 samples presenting high methylation 7 were from Category A cases (58.3%). Consistently, the methylation level distributions assessed in DNA from subjects presenting the CCEF clinical categories A, B and D was not statistically different (Figure S11). Instead, the majority (11/16) of participants in the low methylation group carried Pathogenic, Likely Pathogenic variants or Variants of Uncertain Significance according to ACMG classification^60^ (P/LP/VUS variants) in *SMCHD1* (Fisher exact test p-value 0.0003). This association was confirmed by the comparison of the methylation profile distributions of *SMCHD1* P/LP/VUS variants carriers and non-carriers (Figure 4, Kolmogorov-Smirnov test p-value <= 1e-16). Identical patterns were recovered also when methylation levels on the hg38 assembly were assessed with our approach (Figure 5).

**Figure 6.**
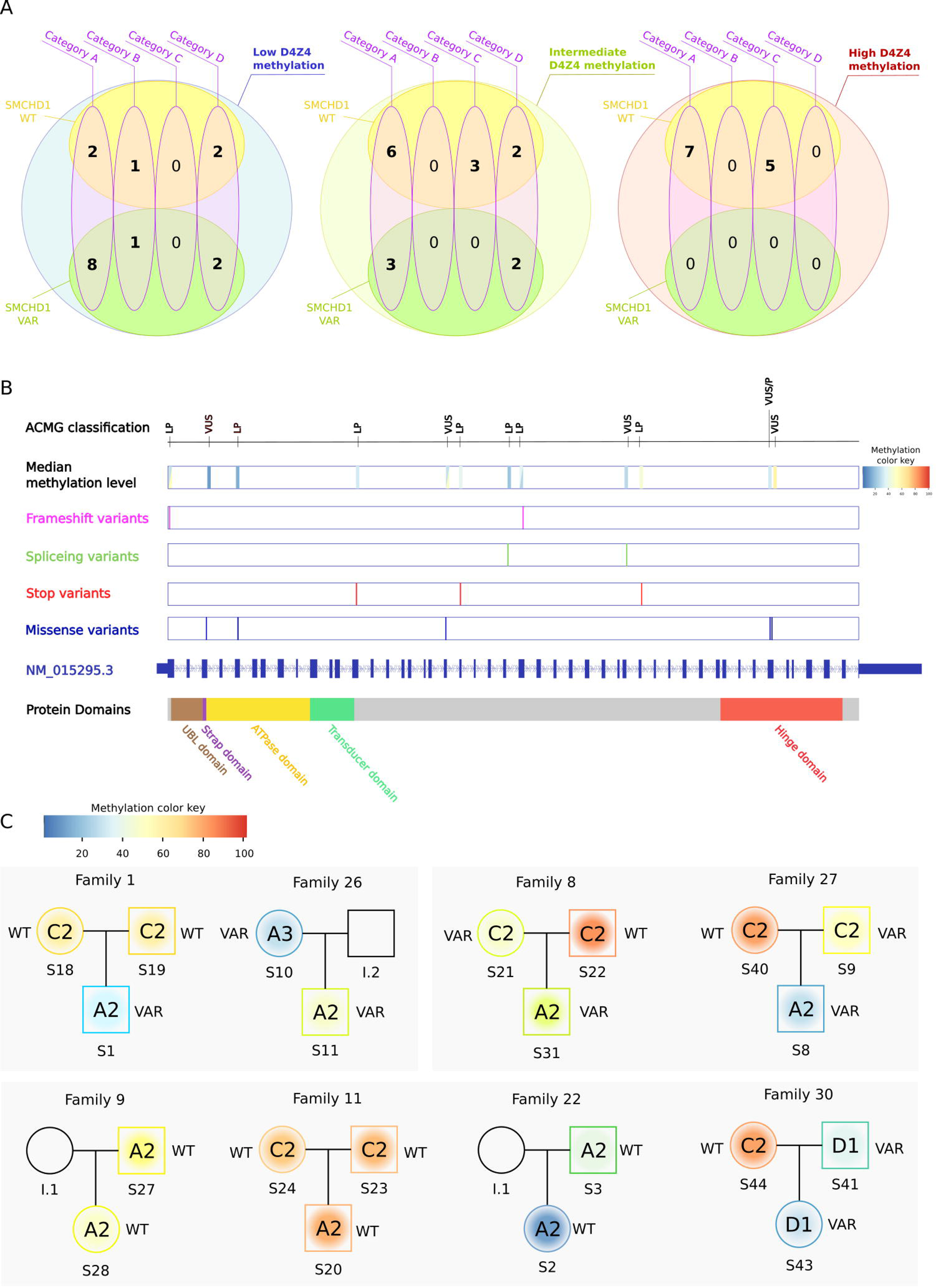
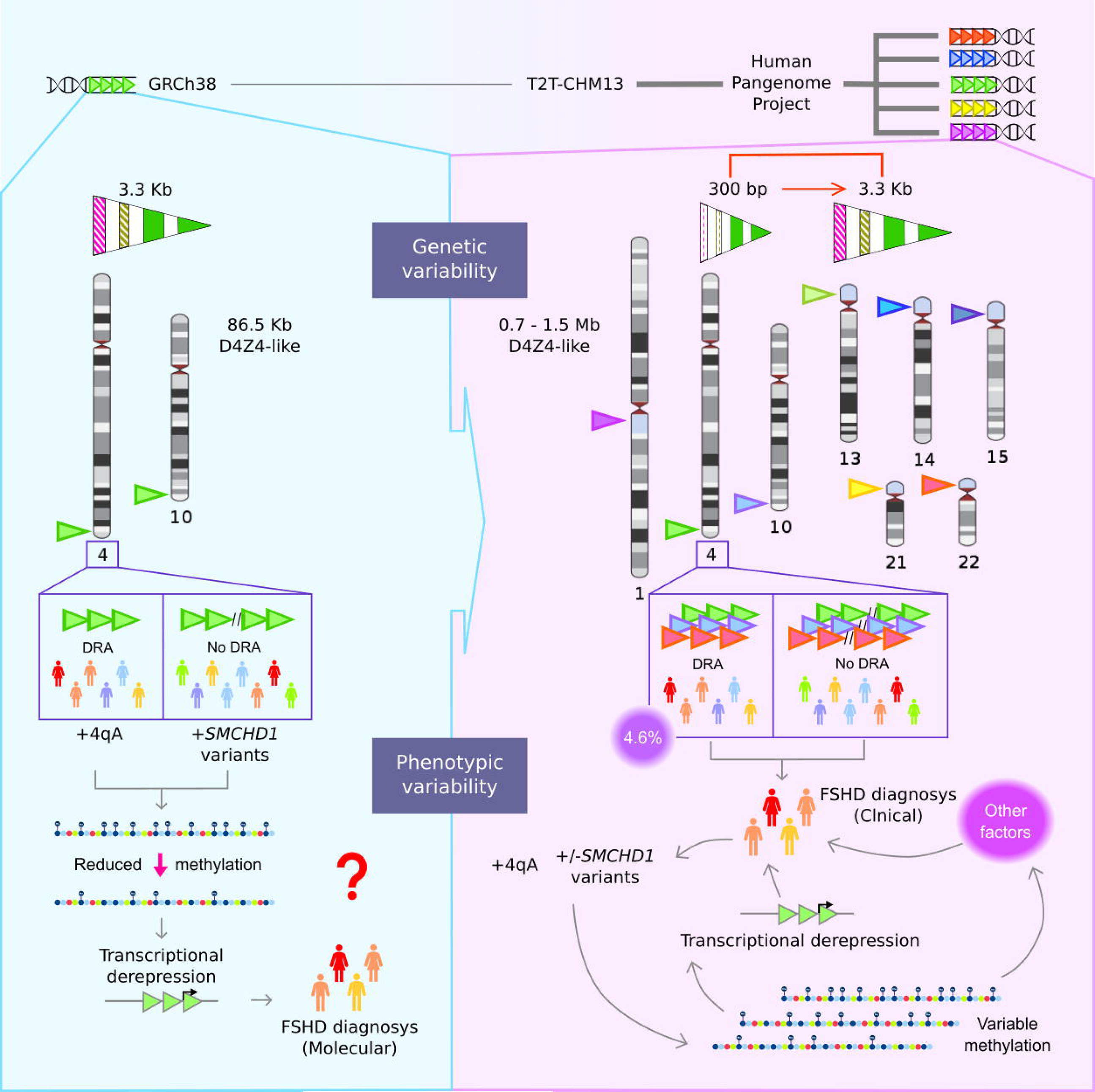
Genotype-phenotype correlation. (A) Venn diagrams reporting the number of subjects presenting different combinations of 4q/10q-specific D4Z4 methylation level, *SMCHD1* mutational status and FSHD clinical category. (B) ACMG classification, median 4q/10q-specific D4Z4 methylation of variant carriers, variant type and location are reported for each *SMCHD1* variant. When the variant is carried by more than one subject, median D4Z4 methylation status of all the subjects is reported. (C) Distribution of 4q/10q-specific D4Z4 methylation, *SMCHD1* mutational status and FSHD clinical category in the 8 trios/parent-child couples included in the study. Circles/squares are colored according to the median 4q/10q-specific D4Z4 methylation level; the color is fading as it represents the variability of CpG methylation pattern in each individual. WT= subject carrying no *SMCHD1* variants, VAR= subject carrying a damaging *SMCHD1* variants.

## Discussion

### The pangenome assemblies provide novel hints to evaluate the significance of the D4Z4-like sequences in the clinical setting

Thirty years ago, reduction below a critical threshold of the number of repeats belonging to the D4Z4 macrosatellite at 4q35 was causally associated with FSHD. D4Z4 is part a family of repetitive elements characteristic of heterochromatin and it was then known that many D4Z4-l were present in the human genome^66^. At that time, the lack of a complete genome assembly hampered the possibility of fully deciphering the significance of molecular and epigenetic findings in FSHD. Very recently, these limitations have been overcome by the availability of complete, high quality haplotype level genome assemblies. By analyzing the haplotypes from the PGR we collected relevant information on the size, the distribution and the composition of the D4Z4 locus, including distal sequence elements that are considered a hallmark of the FSHD molecular signature. We established that the number of 3.3 kb D4Z4 repeats included in each array ranges from 6 RU to 89 RU^40^. We observed that the distance between the qA sequences and the last D4Z4 repeat varies. We also confirmed that qA sequences are more distal compared to qB sequences; the pLam sequence is in association with qA haplotypes^18^, and valid PAS is exclusively associated with 4q^64^; chr 10 D4Z4 repeats were all marked by qA, with a single exception: a 10qB allele carrying 1 RU. Notably, PGR haplotype analysis confirmed the high frequency of the permissive 4qA-PAS haplotype, which is comparable to a common polymorphism^61^. In fact, in 4.6% of cases we identified permissive haplotypes consisting of reduced 4q D4Z4 arrays with ≤ 8 RU and the 4qA-PAS. Since all ter-*DUX4* and pLam sequence elements associated with 4qA alleles were found to be conserved and functionally intact, we speculate that disruptive genomic variants at the ter-*DUX4* are unlikely to account for the staggering discrepancy between the frequency of the presumed causative 4qA-PAS allele and the prevalence of FHSD. Furthermore, no skewed association has been detected between 4q D4Z4 arrays copy number and qA/B alleles^19^, indicating that non-permissive 4qB alleles and potentially permissive 4qA alleles do not have a different selective pressure for the mainteinance of the physiological number of D4Z4 RU.

The consistency between data obtained from the analysis of PGR and previous observations^6,61,66^ shows the validity of the PGR to advance knowledge of the molecular signature used to diagnose FSHD on a world-wide scale.

The PGR analysis also identified 6 haplotypes with peculiar annotations where 2 or more qA/qB probes were assigned to the same chromosome and none to the other chromosome (Table S9). This condition has been reported before^62^ and is ascribable to the spreading of repetitive and polymorphic D4Z4-l at different loci of the genome^63,67^.

Our comparative analyses of complete D4Z4-l (Figure 2C) highlighted that D4Z4 repeats, as well as the ter-*DUX4*, located at chr 4 present a higher variability compared to chr 10. In addition, PGR haplotypes from different ancestries showed different levels of variability in the number of complete D4Z4-l, with a significant difference in D4Z4 copy number between individuals of African ancestry, native Americans and far east Asians.

### D4Z4 methylation assay must consider the T2T sequence for the identification of 4q/10q-specific CpGs

Reduced CpG methylation at D4Z4 has been widely studied and proposed as a biomarker for the presence of FSHD. Previous studies of D4Z4 methylation presented some drawbacks: technical limitations and lack of reproducibility led to discordant results regarding which regions are the most representative for D4Z4 methylation status^20,32,68–74^. In addition, the correlation between D4Z4 methylation level and FSHD clinical status has often been postulated, but different technical settings in the various studies, the lack of clear clinical evaluation of patients and the prominent absence of family studies had hindered the interpretation of results^20,41,73,74^. In the present work, we comprehensively evaluated the D4Z4 methylation status by considering a high number of reads and excluding technical biases introduced by the previous studies based on hg38 and on standard bioinformatics pipelines. We established (Figure S2) that the primer sets BSS-D1 and BSS-D6 identify CpG methylation signals deriving from 4q/10q-specific regions only, whereas the primer sets BSS-D3 and BSS-D5 amplify D4Z4-l distributed not only at 4q and 10q but also on chromosome 1 and on the short arms of acrocentric chromosomes. Thus, the study of D4Z4 CpG methylation can be hazardous if the distribution of D4Z4-l elements and their variability are not correctly taken into account.

Despite all this, the application of short read-based assays for the analysis of repetitive elements of relatively large size has limitations. When non-converted reads were aligned to both hg38 and T2T, more than 50% of mapped reads did not align at expected target regions. Nevertheless, the total fraction of mapped reads and the number of testable CpGs were higher when aligned to T2T compared to hg38, confirming that T2T provides a more accurate representation of the human genome. We can anticipate that long read sequencing technologies, such as Oxford Nanopore, will provide relevant technical advances and open new possibilities for the study of repetitive elements’ organization and function in the genome. In this respect, Butterfield and colleagues (2023)^75^ applied targeted nanopore sequencing to FSHD patients and healthy subjects and showed that an asymmetric methylation gradient forms in a length-dependent manner at the proximal end of the D4Z4 repeat array reaching saturation approximately at the 10th repeat. They also observed a highly similar methylation pattern at 4q and 10q which was irrespective of the clinical phenotype (FSHD or healthy)^75^. This notwithstanding, at present, long-read sequencing technologies are not mature for systematic large-scale applications in the clinical settings. Instead, the targeted CpG methylation analysis we set up is manageable on a large scale as it follows a simple protocol, is less expensive and highly reproducible. Remarkably, our data are consistent with the results obtained by Butterfield and colleagues.

### D4Z4 CpG methylation level is not a predictor of FSHD disease

Beside the formal demonstration that primer design is a crucial point in the analysis of D4Z4 methylation, our study also revealed that contrary to common knowledge^20^, not all DNAs from myopathic patients referred as FSHD2 (26 index cases) presented a low methylation level. Our analysis identified three clusters with low, intermediate and high D4Z4 CpG methylation (mean at 24%, 48% and 71% respectively). Figure 6 depicts the results of several comparisons regarding the following parameters: clinical phenotype, D4Z4 methylation level and *SMCHD1* mutational status. Figure 6A shows that some genotype-phenotype intersections are not in agreement with the current indications for FSHD diagnosis^76,77^. Remarkably, we observed the full range of D4Z4 methylation levels in DNA from the 26 participants presenting a classical FSHD phenotype (CCEF Clinical Category A); of those 16 carriers of a damaging *SMCHD1* variant, 11 presented low 4q/10q-specific D4Z4 CpG methylation, 5 displayed intermediate CpG methylation. We also observed 5 samples with low CpG methylation with no damaging *SMCHD1* or *DNMT3B* or *LRIF1* variants and presenting classical (2), incomplete (1) or complex (2) phenotypes. underlying the genetic heterogeneity of this molecular phenotype.

The variants we identified were distributed all along *SMCHD1* gene body and did not provide information on specific SMCHD1 region/domain that might account for the reduced methylation at 4q/10q-specific D4Z4 sequences (Figure 6B).

To further explore this aspect, we investigated genotype-phenotype correlation in eight trios/parent-child pairs participating to this study (Figure 6C, Table S7). We observed participants presenting the classical FSHD phenotype (CCEF Category A), healthy subjects (CCEF Category C) or complex phenotypes presenting atypical features (CCEF Category D). According to our data, only in families 1 and 26 subjects presenting a classic FSHD phenotype displayed a reduced CpG methylation profile and carried a damaging heterozygous *SMCHD1* variant. In family 22 the classic clinical phenotype, in presence of D4Z4 reduced methylation, was not associated with variants in known chromatin modifiers. In family 8 the probands presented the classic clinical phenotype associated with intermediate D4Z4 methylation level in presence of a damaging *SMCHD1* variant, as the healthy mother. In family 27 the proband presented the Category A phenotype associated with reduced CpG methylation and a damaging *SMCHD1* variant, whereas intermediate D4Z4 methylation level was detected in the healthy father carrying the same *SMCHD1* allele. In family 30 father and daughter presented a complex phenotype with atypical clinical symptoms (Category D1); both reported clinical onset at pelvic girdle and distal lower limbs. They were heterozygous for *SMCHD1* damaging variant and presented reduced D4Z4 CpG methylation. In families 9 both the probands and her father presented Category A phenotype without displaying reduced D4Z4 CpG methylation and carrying WT *SMCHD1* alleles, as the proband in family 11.

Altogether, these results suggest that SMCHD1 modulates, directly or indirectly, D4Z4 methylation at 4q and 10q, at the same time it seems that deleterious variants at *SMCHD1* are not sufficient for the pathogenesis of FSHD and cannot be considered faithful indicators of FSHD clinical status.

In conclusion, our work uncovered the complexity of the genomic setting of D4Z4-l and showed that a molecular diagnostic test for FSHD2 based on *SMCHD1* inactivation and D4Z4 hypomethylation must follow rigorous protocols to avoid biased interpretation. Molecular analysis should be considered together with precise clinical assessment and complemented by family studies for the proper interpretation of results. Moreover, the variability unveiled by our analysis should warn about the risk of relying on a single ideal reference genome sequence in FSHD. In fact, this simplification could strongly narrow our capacity of observing the variability ascribable to each single human genome producing biased interpretations of sequencing data. Finally, our study indicates that a linear approach to diagnose FSHD is obsolete and novel genomic approaches integrated with the precise phenotypic description of patients and their families are needed for the comprehension of the molecular network leading to muscle wasting in FSHD.

## Supporting information

Supplementary Salsi et al MEDRXIV 2023

## Declarations

### Acknowledgements

We are grateful to Francesca Brocco for contributing to the molecular analysis. In addition, we are indebted to all FSHD patients and their families for participating in this study. We acknowledge the Fondazione di Modena for FAR-FOMO 2021 grant funding this work.

### Authors’ contributions

RGT, VS, MC conceived the study concept and supervised the project; VS, SP conducted the molecular analysis; MC conducted bioinformatics analysis; PK and MK contributed to bioinformatic analysis; LR, SCP, MGD, CR, SB, LM, DL contributed to the patients clinical analysis and sample collection; FMS, GP contributed to results discussion; RGT, VS, MC, SP wrote the paper (original draft); all authors read and approved the final manuscript.

### Web sources

The PGR dataset analyzed during the current study is available at https://github.com/human-pangenomics/HPP_Year1_Assemblies.

### Availability of data and materials

Data generated and/or analyzed during the current study and not included in this published article are available from the corresponding author on reasonable request.

### Ethics approval and consent to participate

Signed informed consent was obtained from all the subjects prior to the inclusion in the study. The study was approved by the ethics committee of Emilia Romagna Area Vasta Nord 743/2022/OSS/UNIMO SIRER ID 5111.

