## Supplementary Salsi et al MEDRXIV 2023 for "A human pan-genomic analysis reconfigures the genetic and epigenetic make up of facioscapulohumeral muscular dystrophy"

### Supplemental Figures

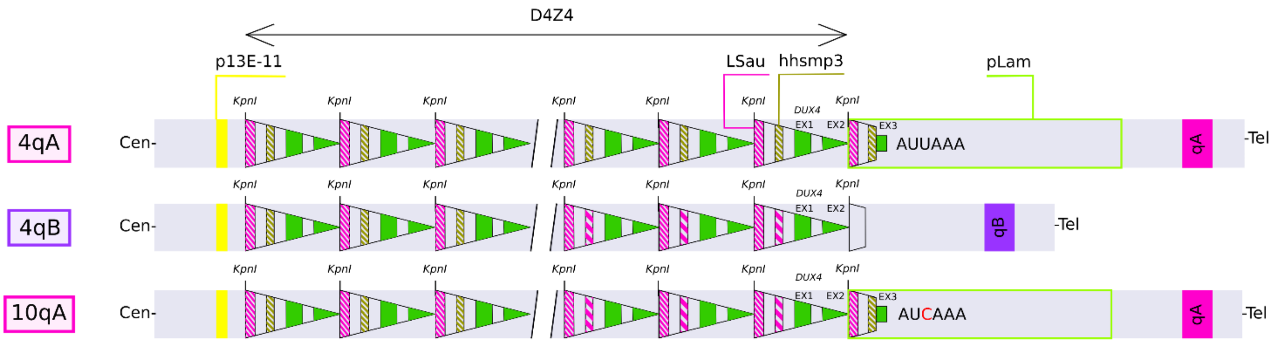

### Figure S1. 4q35 haplotypes

D4Z4 haplotypes include a variable number of D4Z4 tandemly arrayed elements including a *DUX4* ORF (exons 1 and 2) containing a double homeobox and the heterochromatic elements Lsau and hhsmp3. 4q alleles are usually associated to either qA or qB haplotype while 10q alleles are only associated to qB haplotype. qA haplotype includes the pLam sequence carrying an AUUAAA functional polyadenylation signal on chromosome 4q and an AUCAAA non-functional polyadenylation signal on chromosome 10q.

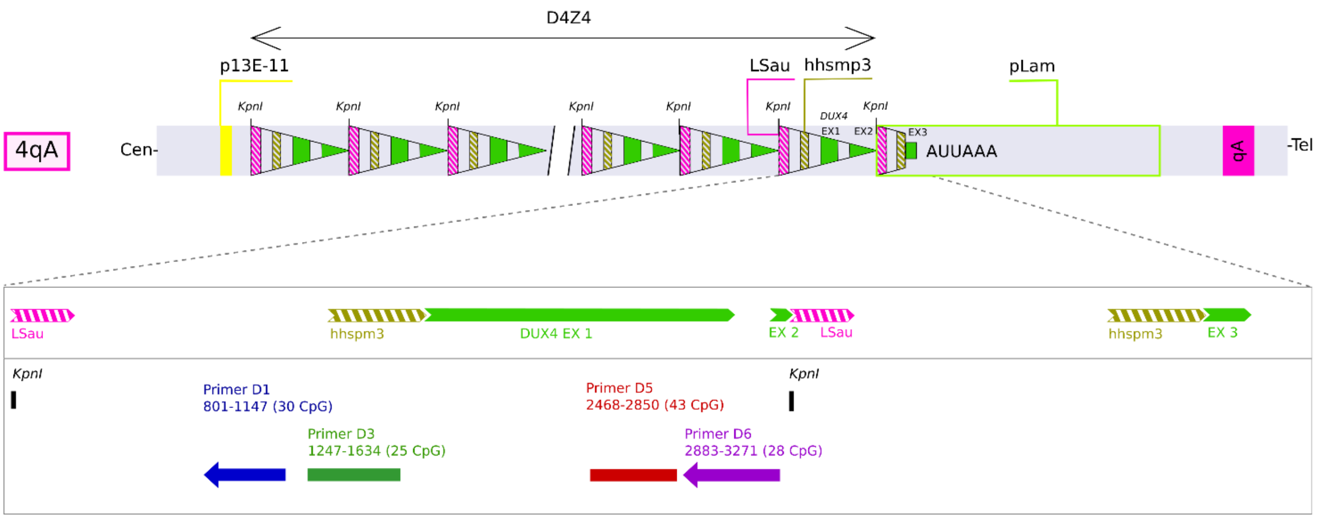

**Figure S2. Amplicon locations at D4Z4 repeats**

The D4Z4 array at 4q35: arrows represent the 3.3 Kb repeats, the p13E-11 probe annealing site is shown in yellow, the pLAM sequence is indicated by a green box. An expanded representation on the last repeat along with the pLam is shown at the bottom. LSau heterochromatic domains and hhspm3 are reported in magenta and brown; *DUX4* exons are reported in green. The position of the amplicons generated from the designed primer sets are reported (coordinates are relative to the 3.3 kb KpnI to KpnI repeat). Primer D1 and Primer D6 were designed on the reverse strand, while Primer D3 and Primer D5 on the forward strand.

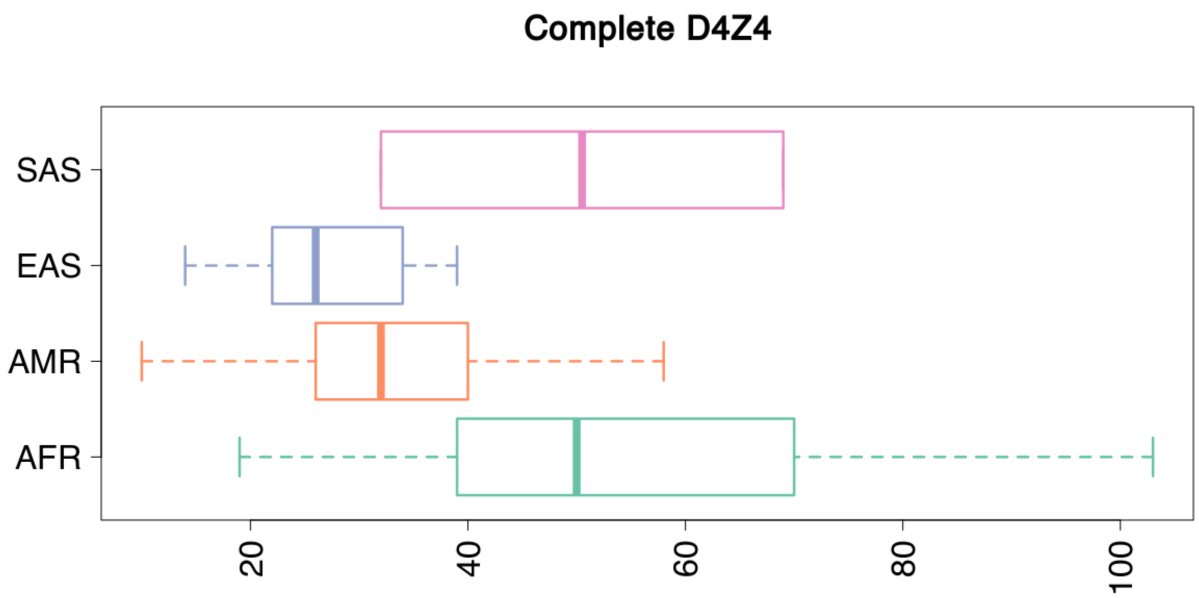

**Figure S3. Estimated copy number of D4Z4 repeats in human geographic macro-populations**

Copy numbers are displayed on the X axis. Distributions of values are represented in the form of a boxplot. Human geographic macro-populations are indicated on the Y axis.

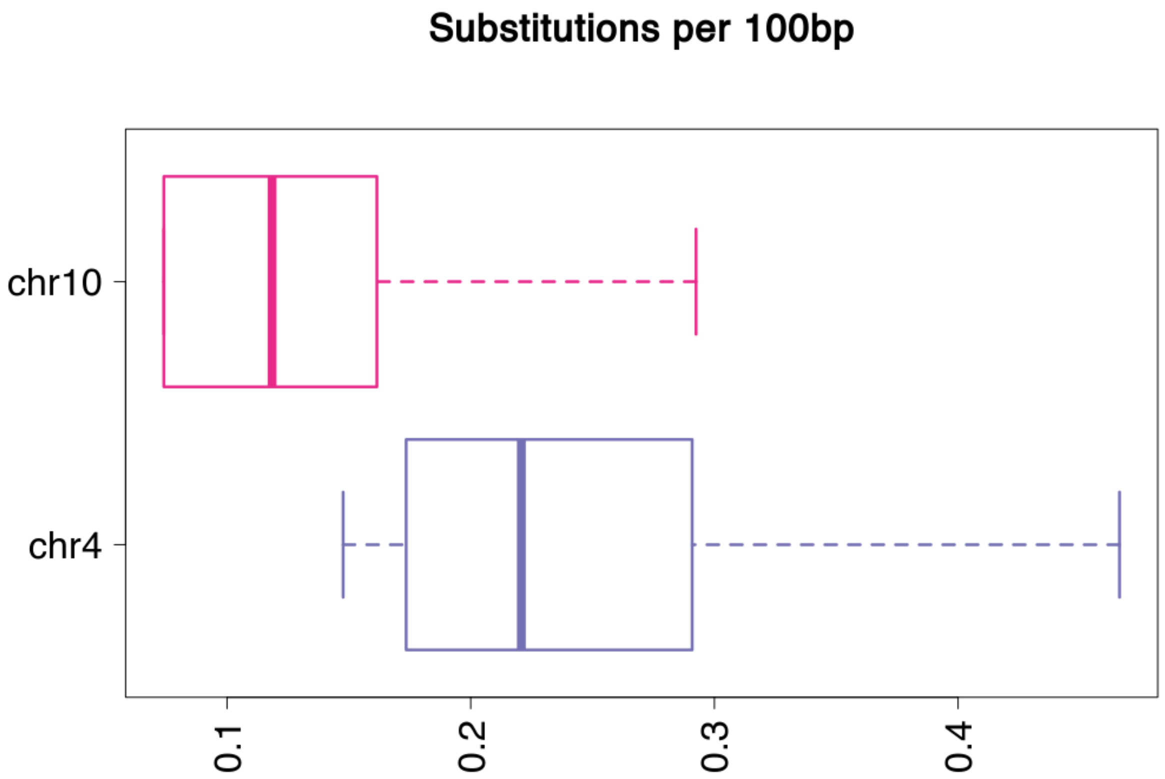

**Figure S4. Estimated nucleotide substitution rates in D4Z4 repeats**

Substitutions rates (substitutions per 100bp) are shown, cumulatively, for repeats assigned to chr 10 and chr 4. Boxplots are used to display the distribution of values. Chromosomes are indicated on the Y axis.

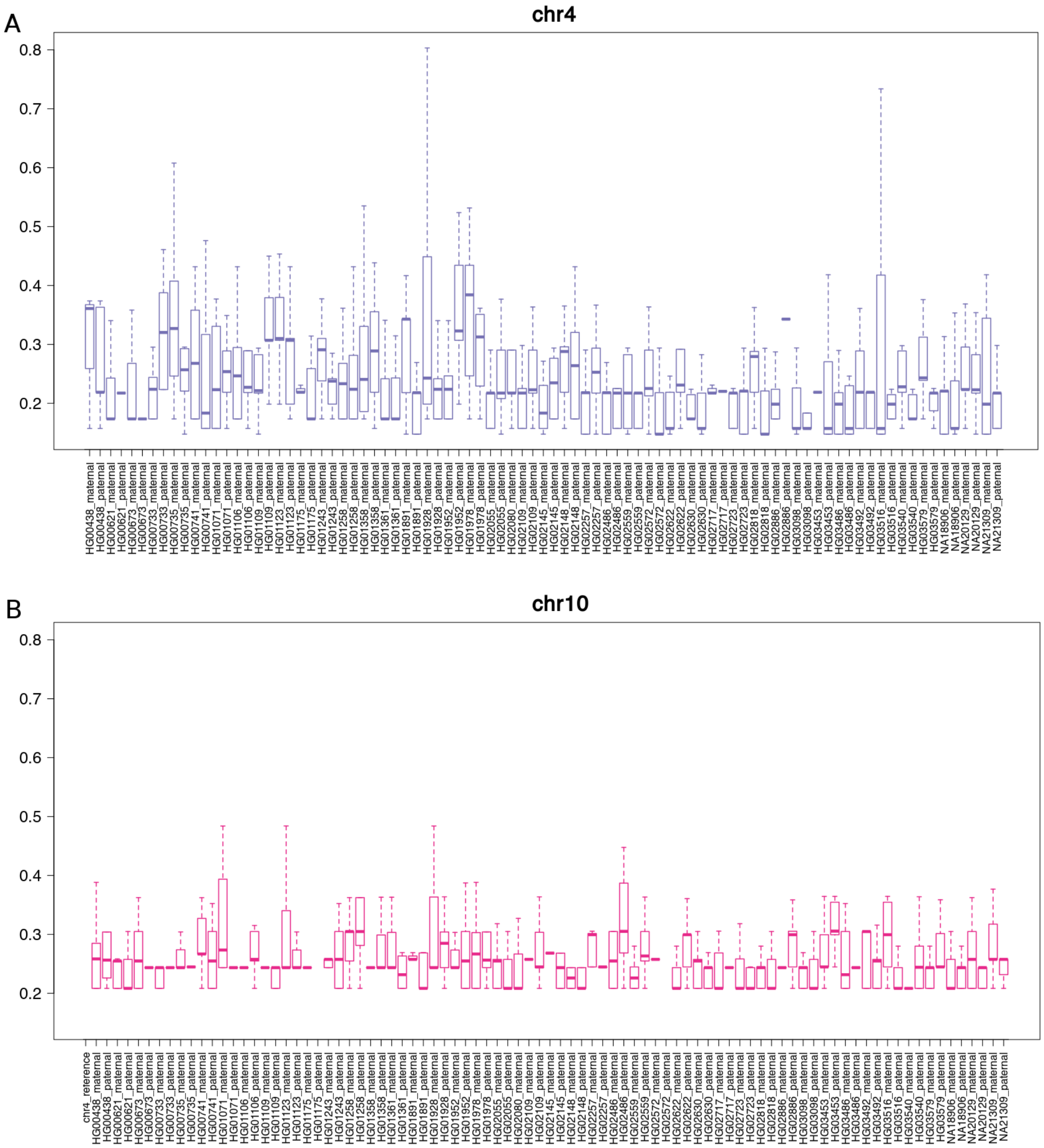

**Figure S5. Estimated nucleotide substitution rates in D4Z4 repeats in haplotypes level assemblies**

Substitutions rates (substitutions per 100bp) are shown, for repeats assigned to chr 10 and chr 4, for every haplotype level assembly from the PGR project. Boxplots are used to display the distribution of values. Haplotypes are reported on the x-axis, substitution rates on the y-axis. (A) chr 4. (B) chr 10.

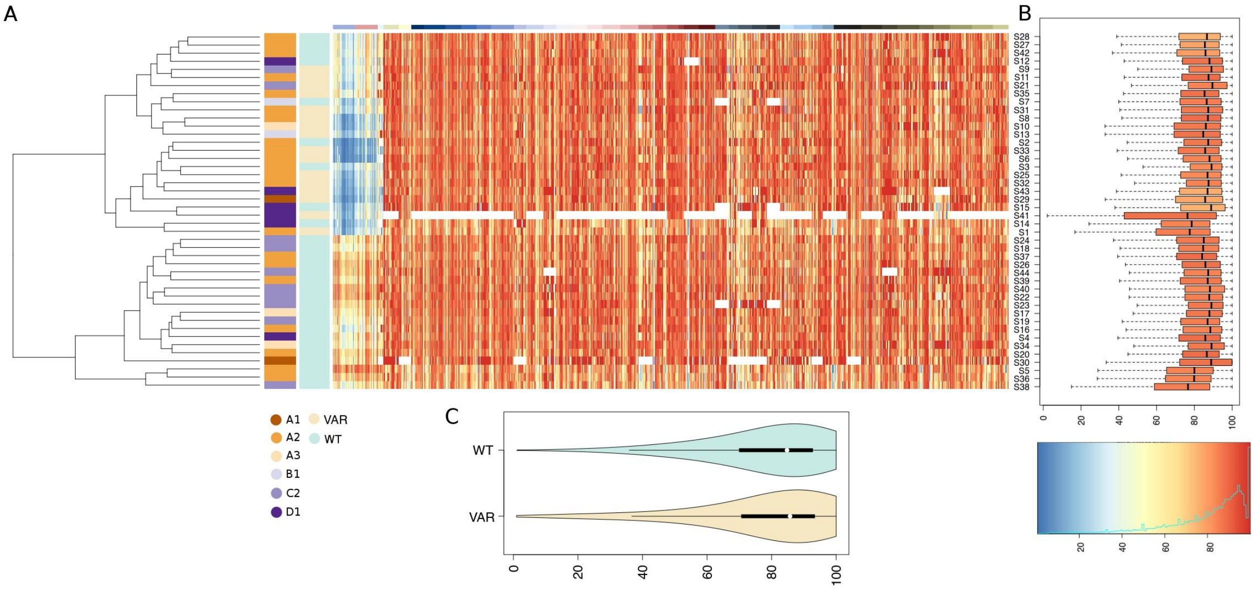

**Figure S6. Cohort subjects' methylation profiles, T2T**

(A) Heatmap of methylation profiles, with individuals reported on the rows, and CpGs on the columns. A total of 938 distinct CpGs are shown. Missing data are indicated in white. The dendrogram shows a clustering of the subjects based on the observed methylation profiles. Colored vertical bars to display the stratification of the individuals according to CCEF grades, and the presence/absence of deleterious genomic variants in *SMCHD1*. Color code legends are displayed directly under each bar. The colored-bar at the top demarcates distinct 95% identity regions. (B) Boxplots display average methylation levels of the 938 CpGs in every subject. (C) Violin-plot of average methylation levels, in subjects with/without deleterious genetic variants in *SMCHD1*.

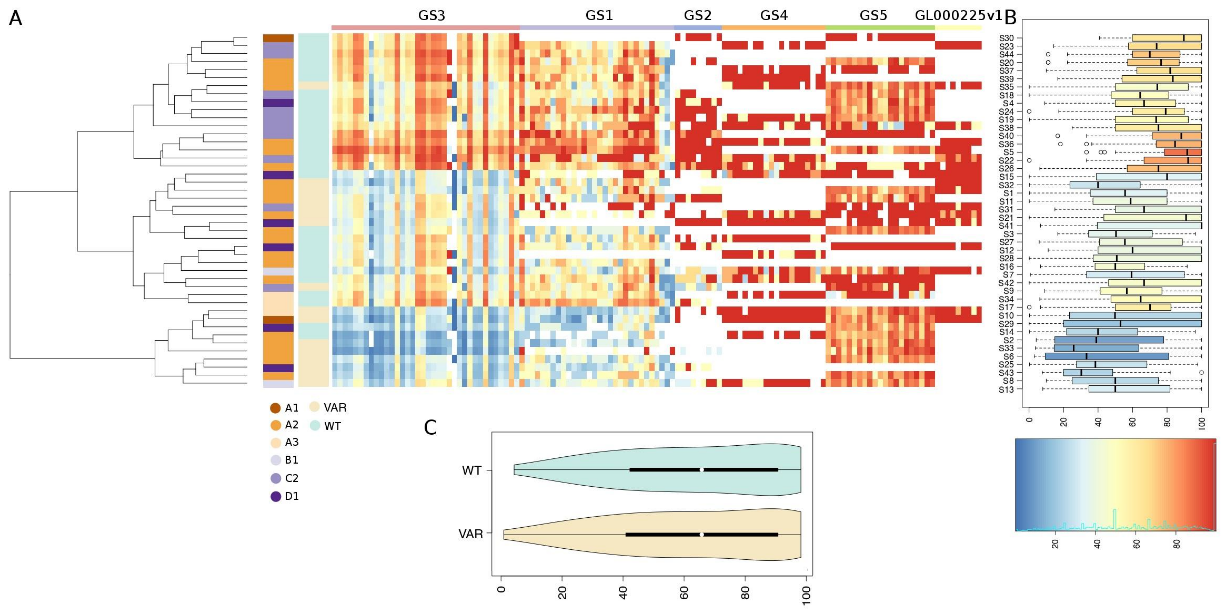

**Figure S7. Cohort subjects' methylation profiles, hg38**

(A) Heatmap of methylation profiles, with individuals reported on the rows, and CpGs on the columns. A total of 125 distinct CpGs are shown. Missing data are indicated in white. The dendrogram shows a clustering of the subjects based on the observed methylation profile. Colored vertical bars display the stratification of the individuals according to CCEF grades, and the presence/absence of deleterious genomic variants in *SMCHD1*. Color code legends are displayed directly under each bar. The colored-bar at the top demarcates distinct 95% identity regions. (B) Boxplots display average methylation levels of the 125 CpGs for every subject. (C) Violin-plot of average methylation levels, in subjects with/without deleterious genetic variants in *SMCHD1*.

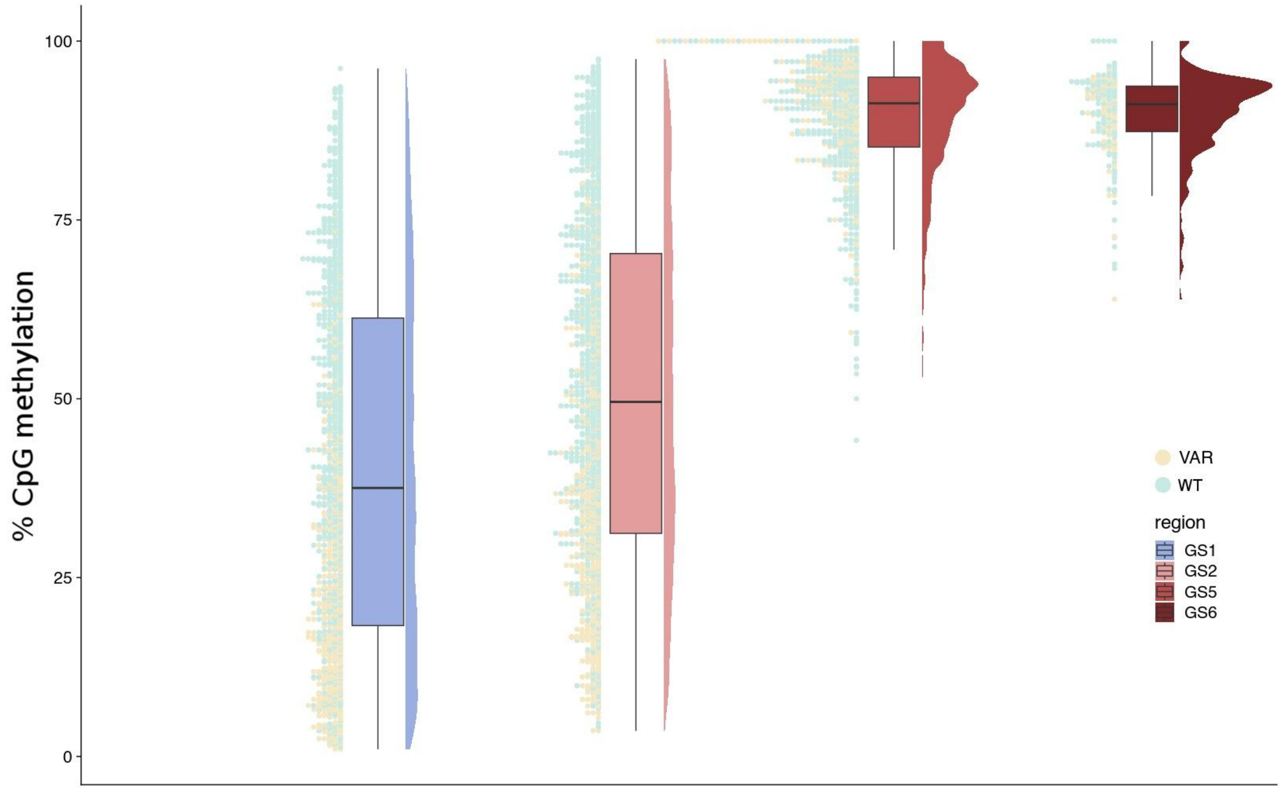

**Figure S8. Raincloud plot of methylation distribution per group of sequences (GS)**

Raincloud plot of D4Z4 CpGs methylation level distribution for the GS1 (Primer D6), GS2 (Primer D1), GS5 and GS6 consensus sequences. Dots on the left of each plot represent methylation levels at single CpG resolution and are colored according to the *SMCHD1* mutational status of the corresponding subject.

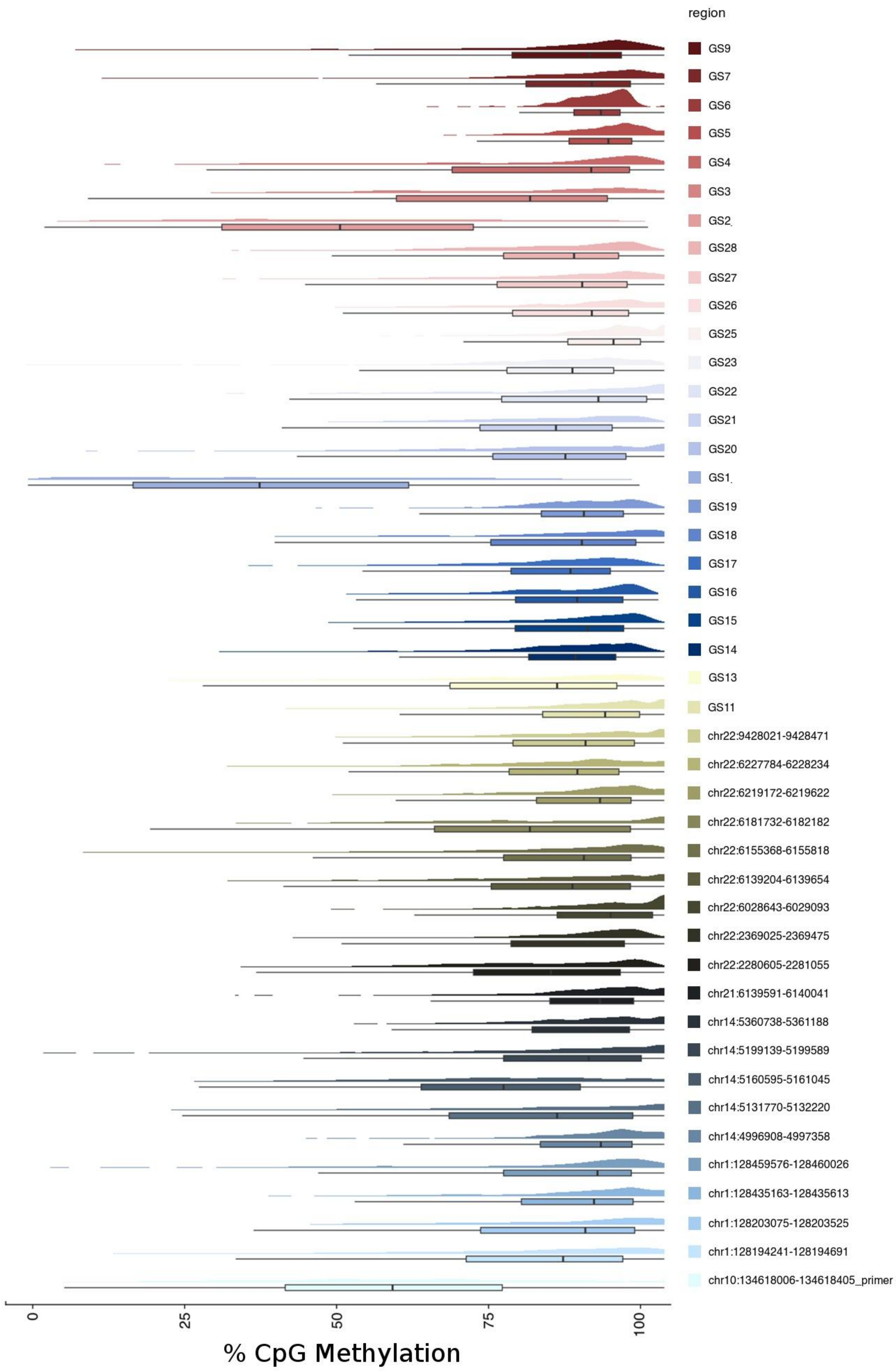

**Figure S9. Raincloud plot of CpG methylation at all group of sequences (GS)**

Raincloud plot of D4Z4 CpGs methylation distribution for all GSs.

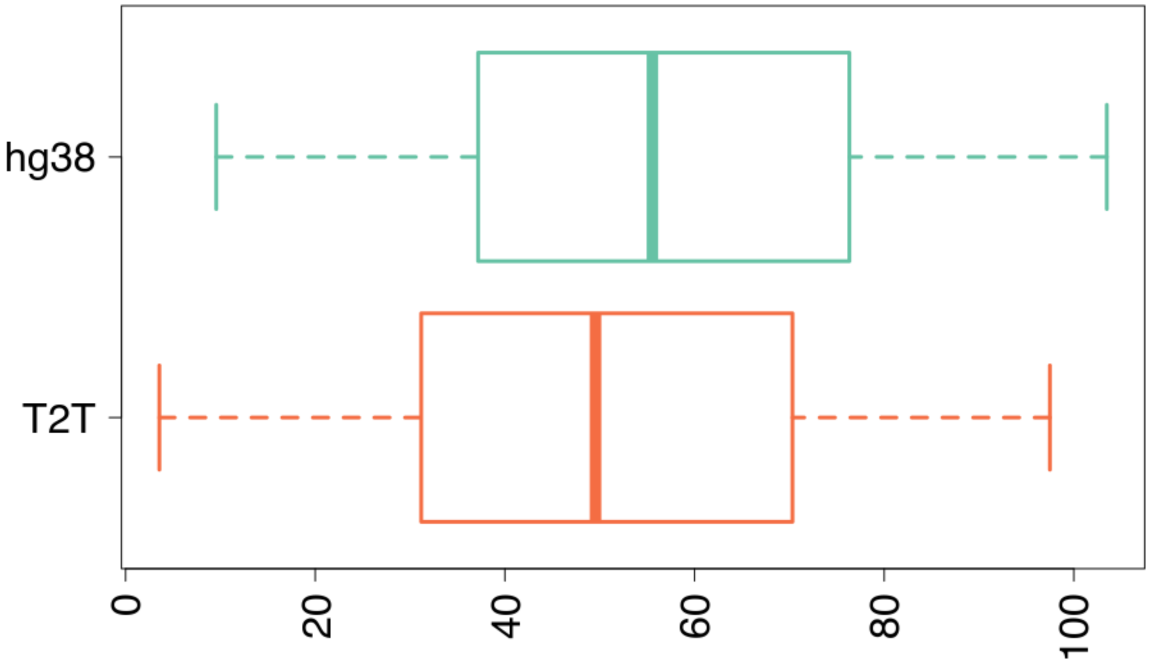

**Figure S10. Average methylation level at Primer D1 on T2T and hg38**

The distributions of values are displayed in the form of a boxplot. Genome assemblies are indicated on the Y axis, average methylation levels on the X-axis.

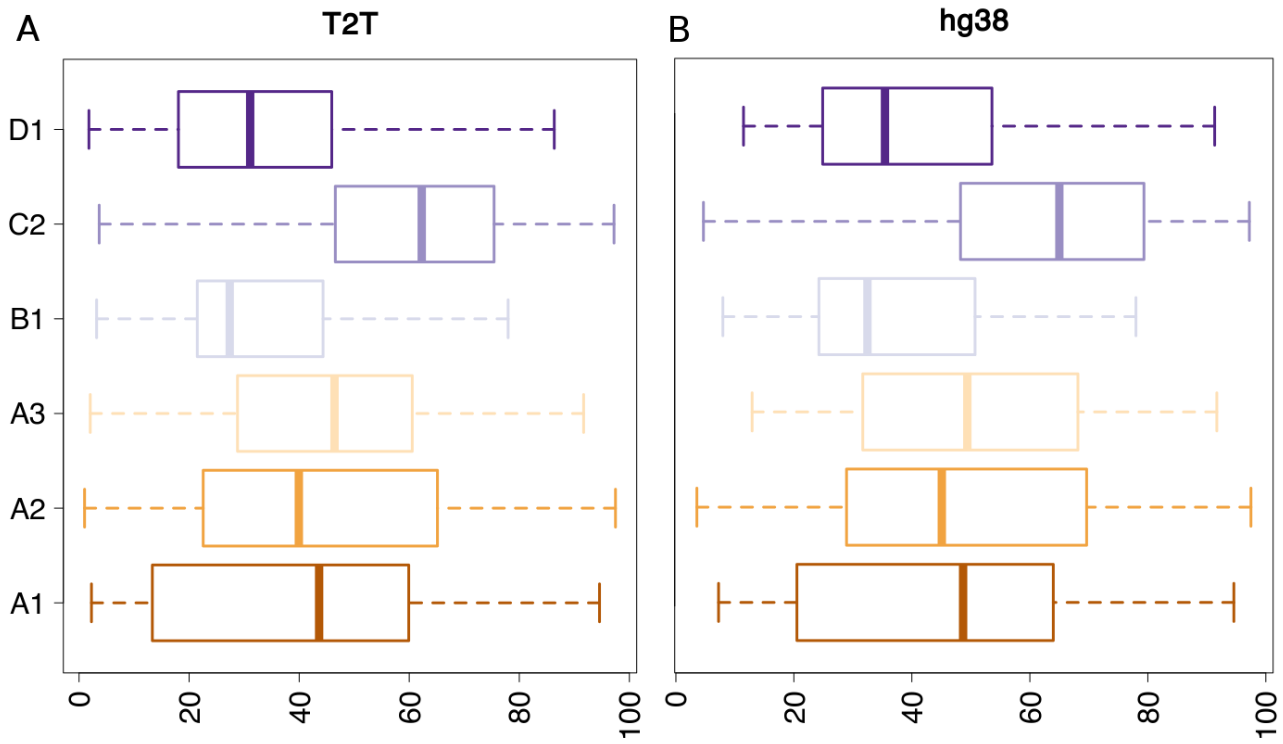

**Figure S11. Average methylation level at primers targeted CpGs, according to CCEF grade**

(A) T2T reference assembly. (B) hg38 reference assembly. Boxplots are used to display the distribution of values. CCEF grades are reported on the Y axis, average methylation levels on the X-axis.

### Supplemental Tables

#### Table S1. Distribution of subjects according to sex and FSHD category

| | | Sex | | FSHD Category | | | | | |
|---|---|---|---|---|---|---|---|---|---|
| | N° subjects | Male | Female | A1 | A2 | A3 | B1 | C2 | D1 |
| **FSHD2** | 26 | 18 | 8 | 2 | 14 | 3 | 2 | - | 5 |
| **Relatives** | 14 | 7 | 7 | - | 3 | - | - | 10 | 1 |
| **FSHD1** | 3 | 2 | 1 | - | 3 | - | - | - | - |
| **Relatives** | 1 | - | 1 | - | 1 | - | - | - | - |
| **TOT** | 44 | 27 | 17 | 2 | 21 | 3 | 2 | 10 | 6 |

#### Table S2. Forward (FW) and reverse (RW) sequence of primers

| Primer name | FW | RW |
|---|---|---|
| **D1** | 5’- AGA CCC CAG AGA GGA GAT GCC T-3’ | 5’-CCC AGG CTG AGC CCT GCA A-3’ |
| **D3** | 5’- GAG AGA GGG CCT GGC ACA CTC AAG A -3’ | 5’- GGC AGG CAG GCT CCA CCC CTT CAT -3’ |
| **D5** | 5’- GGC AGA GGG GGT CTC CCA ACC TGC -3’ | 5’- GGA TGC CTT GCA TCT GCC CCT GCC -3’ |
| **D6** | 5’- GGA TGC CCA GGA AAG AAT GGC AGT T-3’ | 5’- CCC TGG TCT GCA CTC CCC TG -3’ |
| **BSS-D1** | 5’- AGA TTT TAG AGA GGA GAT GTT T-3’ | 5’-CCC AAA CTA AAC CCT ACA A-3’ |
| **BSS-D3** | 5’- GAG AGA GGG TTT GGT ATA TTT AAG A -3’ | 5’- AAC AAA CAA ACT CCA CCC CTT CAT A -3’ |
| **BSS-D5** | 5’- GGT AGA GGG GGT TTT TTA ATT TGT -3’ | 5’- AAA TAC CTT ACA TCT ACC CCT ACC -3’ |
| **BSS-D6** | 5’- GGA TGT TTA GGA AAG AAT GGT AGT T-3’ | 5’- CCC TAA TCT ACA CTC CCC TA -3’ |

D= non-converted sequence; BSS-D= BSC sequence

### Table S3. Amplicon size and number of analyzed CpGs

| Primer couple | Amplicon size | N° CpGs |
|---|---|---|
| **BSS-D1/D1** | 347 bp | 30 |
| **BSS-D3/D3** | 388 bp | 25 |
| **BSS-D5/D5** | 383 bp | 43 |
| **BSS-D6/D6** | 389 bp | 28 |
| **Total** | 1507 bp | 126 |

**Table S4. Primers target region**

| chr | T2T_notConverted | T2T_converted | hg38_notConverted | hg38_converted |
|---|---|---|---|---|
| chr4 | 134 | 118 | 52 | 39 |
| chr10 | 139 | 140 | 37 | 53 |
| chr11 | 0 | 3 | 0 | 5 |
| chr12 | 0 | 4 | 0 | 2 |
| chr13 | 36 | 22 | 0 | 4 |
| chr14 | 170 | 105 | 4 | 3 |
| chr15 | 70 | 40 | 0 | 3 |
| chr16 | 0 | 2 | 0 | 5 |
| chr17 | 0 | 3 | 0 | 3 |
| chr18 | 0 | 3 | 0 | 4 |
| chr1 | 73 | 39 | 0 | 5 |
| chr20 | 0 | 2 | 0 | 2 |
| chr21 | 92 | 48 | 0 | 0 |
| chr22 | 166 | 63 | 0 | 0 |
| chr2 | 0 | 4 | 0 | 5 |
| chr3 | 2 | 4 | 0 | 4 |
| chr5 | 2 | 2 | 0 | 4 |
| chr6 | 0 | 2 | 0 | 2 |
| chr7 | 0 | 2 | 0 | 3 |
| chr8 | 0 | 3 | 0 | 3 |
| chr9 | 0 | 4 | 0 | 5 |
| chrX | 0 | 3 | 0 | 4 |
| chrY | 4 | 0 | 4 | 0 |
| **Tot** | 888 | 616 | 97 | 158 |
| **chr4/chr10** | 273 | 258 | 89 | 92 |

chr: chromosome. T2T_notConverted: total number of genomic regions targeted by non-converted primers in the T2T genome assembly. T2T_converted: total number of genomic regions targeted by BSC primers in the T2T genome assembly. hg38_notConverted: total number of genomic regions targeted by non-converted primers in the hg38 genome assembly. hg38_converted: total number of regions by BSC primers in the hg38 genome assembly.

**Supplemental Material and Methods**

**FASTA reference sequences used for characterization of D4Z4 repeats and 4q35 haplotypes.**

**>D4Z4 pLAM**

GACGCGGGGTTGGGACGGGGTCGGGTGGTTCGGGGCAGGGCGGTGGCCTCTCTTT
CGCGGGGAACACCTGGCTGGCTACGGAGGGGCGTGTCTCCGCCCCGCCCCCTCCA
CCGGGCTGACCGGCCTGGGATTCCTGCCTTCTAGGTCTAGGCCCGGTGAGAGACTC
CACACCGCGGAGAACTGCCATTCTTTCCTGGGCATCCCGGGGATCCCAGAGCCGGC
CCAGGTACCAGCAGGTGGGCCGCCTACTGCGCACGCGCGGGTTTGCGGGCAGCCG
CCTGGGCTGTGGGAGCAGCCCGGGCAGAGCTCTCCTGCCTCTCCACCAGCCCACCC
CGCCGCCTGACCGCCCCCTCCCCACCCCCACCCCCCACCCCCGGAAAACGCGTCGT
CCCCTGGGCTGGGTGGAGACCCCCGTCCCGCGAAACACCGGGCCCCGCGCAGCGT
CCGGGCCTGACACCGCTCCGGCGGCTCGCCTCCTCTGCGCCCCCGCGCCACCGTC
GCCCGCCCGCCCGGGCCCCTGCAGCCTCCCAGCTGCCAGCACGGAGCGCCTGGCG
GTCAAAAGCATACCTCTGTCTGTCTTTGCCCGCTTCCTGGCTAGACCTGCGCGCAGT
GCGCACCCCGGCTGACGTGCAAGGGAGCTCGCTGGCCTCTCTGTGCCCTTGTTCTT
CCGTGAAATTCTGGCTGAATGTCTCCCCCCACCTTCCGACGCTGTCTAGGCAAACCT
GGATTAGAGTTACATCTCCTGGATGATTAGTTCAGAGATATATTAAAA

**>qAprobe_EF71075913**

TTTCATTTAGGAAACAGCTATGACCATGATTACGCCAAGCTTGGTACCGAGCTCGGATC
CACTAGTAACGGCCGCCAGTGTGCTGGAATTCGCCCTTGGCTGCAGAAGCAGCTTCT
CCTCTATGTTCTTCACTGCCTCATACTGTTGTTGACCTTGAAACCTTCTTTTGGTCTAGT
TTTATCAACAGAGCTAGTATTTACATGAGGTTCTACTACATACCAGGTTCCAGAAAGCTA
AATGCTTTTTGTTTGTTTTTATTCACTAAATACAAATCACAACTCTGTTCTCATTACACAC
ACAACAAAATTTAGCTGAGGGAGATTGAGTGACTTTCCCAGGGTCACACAGCTACTAA
TAGCAGAGTAGTGTTTAGATTCATATGGGAATACTGAACACAGAAATGAACCAATGGAA
ACATCCTACGTTCCAAAAGCCTACTCAAGCCATTTGTTCTTATTTTAAGGAAAATATGCT
AATTTTAAACTCCAACTATGTATGAATTTATAGGCATGCGGTAAACTCTAATTGCCACGG
GTGAATATGCAGTAAGAAATTCAGTTCAGTAGATGAAACTCAAAAAAGATTCCGCAGCG
AAAGATCTGAGTTCTACCAAACATACAGAGAAGTAGAAATAGATCATTTCAGAATTTTGG
AGCATCGTTATCATCCAAAATAGGCATAGCATCGACTCAAAGGCGTCGATACTCGCAGT
TAAATTCTTCTCTCCGGGGGGGTAAATTGCAAATAAACATTCGTGTTCGCCGTTATTCC
TTACCCATAA

**>qBprobe_EF71076933**

TAATCCCATAAAGGTCATGGGCCATTTGCCACCTGAACAGTCAGTAACACATGAGTGG
AAAGAAACTGAACACCCAGGGACACCAGAGACCGTTCACTGTAGAGGAAGGAGGCA
GGTATAACTCATAAGGTGATCTTCCTCCAGTCCCAGAGTTGTTCCCCTCTCCTTAAATG
TGGGTCCCATGGAATTCAGACTAGGAAGTAAAGCAAATGAGAAAGGCCTACAGGGGA

GCAGTTCAAATGTGTGGAAAAGGATAGAGCAGCCCCAATGAGGAAGGAAGGCTGGAC
AAGCAATGGATTTGTAGGGAAGACAATGTGCACCCATCGGAGCTCTGATTCCTTCATTT
TCACTACCCTCCCCTGCCCACTAACATAAAAAAAGTATTGATGGCAAGTGTTGGGCCAT
AAGACATTTCCTTGCTTTATAATCTGGATTTGGGGGGTTACATTTCAGAGATAATGAAAA
TCTCCTCCTTTAGTTAACTTTTAATTTCATAATTGTAACTTTGTTTTTACAGATTTGTGAAC
ATGTAAAACAAAGAAAACAGCATGGGAGTAATTTATAATCAACAAATATGTGTTCACTGA
GTGACACTCACATGGCATATGGCATATGGGCATCCTGAGAGTAGGATCGTGGGGCATC
CGATGGTGATTGTCCTTAGAGTTGGTCGGAGCCCAGCCTTCCACGGTCTGCGCACTG
CTGTGTGTACAATCGGTGCCTTCTTTAAGTTCACAGCAATGCCACAAGGCAAATAGCA
CTGTCTTCACTTTCTGTGTGAGAGAACTAAGGCTGGGTGAGAGTATGCAGAGCTGGAT
CATGGATTTGTTTGGCTC

**>D4Z4 sequence KpnI-KpnI 3299bp**

GGTACCAGCAGGTGGGCCGCCTACTGCGCACGCGCGGGTTTGCGGGCAGCCGCCT
GGGCTGTGGGAGCAGCCCGGGCAGAGCTCTCCTGCCTCTCCACCAGCCCACCCCG
CCGCCTGACCGCCCCATCCCCACCCCCCACCCCCCACCCCCGGAAAACGCGTCGTC
CCCTGGGCTGGGTGGAGACCCCCGTCCCGCGAAACACCGGGCCCCGCGCAGCGTC
CGGGCCTGACACCGCTCCGGCGGCTCGCCTCCTCCCTGTCGCCCCCGGGCCACCG
TCGCCCGCCGCCCGGGCCCCTGCAGCCGCCCAGCTGCCAGCACGGAGCGCCTGGC
GGCGGAACGCAGACCCCAGGCCCGGCGCACACCGGGACGCTGAGCGTTCCAGGCG
GGAGGGAAGGCGGGCAGAGATGGAGAGAGGAACGGGAGACCTAGAGGGGCGGAAG
GATGGGCGGAGGGACGTTAGGAGGGAGGCAGGGAGGCAGGGAGGCAGGGAGGAA
CGGAGGGAGAGACAGAGCGACGCAGGGACTGGGGGCGGGCGGGAGGGAGCCGGG
GACGGACGGGGGGAGGAAGGCAGGGAGGAAAAGCGGTCCTCGGCCTCCGGGAGTA
GCGGGACCCCCGCCCTCCGGGAAAACGGTCAGCGTCCGGCGCGGGCTGAGGGCTG
GGCCCACAGCCGCCGCGCCGGCCGGCGGGGCACCACCCATTCGCCCCGGTTCCGG
GGCCCAGGGAGTGGGCGGTTTCCTCCGGGACAAAAGACCGGGACTCGGGTTGCCG
TCGGGTTTTCACCCGCGCGGTTCACAGACCGCACATCCCCAGGCTGAGCCCTGCAA
CGCGGCGCGAGGCCGACAGACCCGGCCACGGAGGAGCCACACGCAGGACGACGG
AGGCGTGATTTTGGTTTCCGCGTGGCTTTGCCCTCCGCAAGGCGGCCTGTTGCTCAC
GTCTCTCCGGCCCCCGAAAGGCTGGCCATGCCGACTGTTTGCTCCCGGAGCTCTGC
GGGCACCCGGAAACATGCAGGGAAGGGTGCAAGCCCGGCATGGTGCCTTCGCTCTC
CTTGCCAGGTTCCAAACCGGCCACACTGCAGACTCCCCCACGTTGCCGCACGCGGG
AATCCATCGTCAGGCCATCACGCCGGGGAGGCATCTCCTCTCTGGGGTCTCGCTCTG
GTCTTCTACGTGGAAATGAACGAGAGCCACACGCCTGCGTGTGCGAGACCGTCCCG
GCAACGGCGACGCCCACAGGCATTGCCTCCTTCACGGAGAGAGGGCCTGGCACACT
CAAGACTCCCACGGAGGTTCAGTTCCACACTCCCCTCCACCCTCCCAGGCTGGTTTC
TCCCTGCTGCCGACGCGTGGGAGCCCAGAGAGCGGCTTCCCGTTCCCGCGGGATCC
CTGGAGAGGTCCGGAGAGCCGGCCCCCGAAACGCGCCCCCCTCCCCCCTCCCCCC
TCTCCCCCTTCCTCTTCGTCTCTCCGGCCCCACCACCACCACCGCCACCACGCCCTC
CCCCCCCCCCCCCCCCCCCCACCACCACCACCCCGCCGGCCGGCCCCAGGCCTCG
ACGCCCTGGGTCCCTTCCGGGGTGGGGCGGGCTGTCCCAGGGGGGCTCACCGCCA
TTCATGAAGGGGTGGAGCCTGCCTGCCTGTGGGCCTTTACAAGGGCGGCTGGCTGG
CTGGCTGGCTGTCCGGGCAGGCCTCCTGGCTGCACCTGCCGCAGTGCACAGTCCG
GCTGAGGTGCACGGGAGCCCGCCGGCCTCTCTCTGCCCGCGTCCGTCCGTGAAATT
CCGGCCGGGGCTCACCGCGATGGCCCTCCCGACACCCTCGGACAGCACCCTCCCC

GCGGAAGCCCGGGGACGAGGACGGCGACGGAGACTCGTTTGGACCCCGAGCCAAA
GCGAGGCCCTGCGAGCCTGCTTTGAGCGGAACCCGTACCCGGGCATCGCCACCAGA
GAACGGCTGGCCCAGGCCATCGGCATTCCGGAGCCCAGGGTCCAGATTTGGTTTCA
GAATGAGAGGTCACGCCAGCTGAGGCAGCACCGGCGGGAATCTCGGCCCTGGCCC
GGGAGACGCGGCCCGCCAGAAGGCCGGCGAAAGCGGACCGCCGTCACCGGATCCC
AGACCGCCCTGCTCCTCCGAGCCTTTGAGAAGGATCGCTTTCCAGGCATCGCCGCCC
GGGAGGAGCTGGCCAGAGAGACGGGCCTCCCGGAGTCCAGGATTCAGATCTGGTTT
CAGAATCGAAGGGCCAGGCACCCGGGACAGGGTGGCAGGGCGCCCGCGCAGGCAG
GCGGCCTGTGCAGCGCGGCCCCCGGCGGGGGTCACCCTGCTCCCTCGTGGGTCGC
CTTCGCCCACACCGGCGAGTGGGGAACGGGGCTTCCCGCACCCCACGTGCCCTGC
GCGCCTGGGGCTCTCCCACAGGGGGCTTTCGTGAGCCAGGCAGCGAGGGCCGCCC
CCGCGCTGCAGCCCAGCCAGGCCGCGACGGCAGAGGGGGTCTCCCAACCTGCCCC
GGCGCGCGGGGATTTCGCCTACGCCGCCCCGGCTCCTCCGGACGGGGCGCTCTCC
CACCCTCAGGCTCCTCGGTGGCCTCCGCACCCGGGCAAAAGCCGGGAGGACCGGG
ACCCGCAGCGCGACGGCCTGCCGGGCCCCTGCGCGGTGGCACAGCCTGGGCCCG
CTCAAGCGGGGCCGCAGGGCCAAGGGGTGCTTGCGCCACCCACGTCCCAGGGGAG
TCCGTGGTGGGGCTGGGGCCGGGGTCCCCAGGTCGCCGGGGCGGCGTGGGAACC
CCAAGCCGGGGCAGCTCCACCTCCCCAGCCCGCGCCCCCGGACGCCTCCGCCTCC
GCGCGGCAGGGGCAGATGCAAGGCATCCCGGCGCCCTCCCAGGCGCTCCAGGAGC
CGGCGCCCTGGTCTGCACTCCCCTGCGGCCTGCTGCTGGATGAGCTCCTGGCGAGC
CCGGAGTTTCTGCAGCAGGCGCAACCTCTCCTAGAAACGGAGGCCCCGGGGGAGCT
GGAGGCCTCGGAAGAGGCCGCCTCGCTGGAAGCACCCCTCAGCGAGGAAGAATACC
GGGCTCTGCTGGAGGAGCTTTAGGACGCGGGGTTGGGACGGGGTCGGGTGGTTCG
GGGCAGGGCGGTGGCCTCTCTTTCGCGGGGAACACCTGGCTGGCTACGGAGGGGC
GTGTCTCCGCCCCGCCCCCTCCACCGGGCTGACCGGCCTGGGATTCCTGCCTTCTA
GGTCTAGGCCCGGTGAGAGACTCCACACCGCGGAGAACTGCCATTCTTTCCTGGGCA
TCCCGGGGATCCCAGAGCCGGCCCAGGTACC
